# A Biomarker-Centric Framework for the Prediction of Future Chronic Pain

**DOI:** 10.1101/2024.04.19.24306101

**Authors:** Matt Fillingim, Christophe Tanguay-Sabourin, Marc Parisien, Azin Zare, Gianluca V. Guglietti, Jax Norman, Bogdan Petre, Andrey Bortsov, Mark Ware, Jordi Perez, Mathieu Roy, Luda Diatchenko, Etienne Vachon-Presseau

## Abstract

Chronic pain is a multifactorial condition presenting significant diagnostic and prognostic challenges. Biomarkers for the classification and the prediction of chronic pain are therefore critically needed. In this multi-dataset study of over 523,000 participants, we applied machine learning to multi-dimensional biological data from the UK Biobank to identify biomarkers for 35 medical conditions associated with pain (e.g., clinical diagnosis of rheumatoid arthritis, fibromyalgia, stroke, gout, etc.) or self-reported chronic pain (e.g., back pain, knee pain, etc). Biomarkers derived from blood immunoassays, brain and bone imaging, and genetics were effective in predicting medical conditions associated with chronic pain (area under the curve (AUC) 0.62-0.87) but not self-reported pain (AUC 0.50-0.62). Among the biomarkers identified was a composite blood-based signature that predicted the onset of various medical conditions approximately nine years in advance (AUC 0.59-0.72). Notably, all biomarkers worked in synergy with psychosocial factors, accurately predicting both medical conditions (AUC 0.69– 0.91) and self-report pain (AUC 0.71–0.92). Over a period of 15 years, individuals scoring high on both biomarkers and psychosocial risk factors had twice the cumulative incidence of diagnoses for pain-associated medical conditions (Hazard Ratio (HR): 2.26) compared to individuals scoring high on biomarkers but low on psychosocial risk factors (HR: 1.06). In summary, we identified various biomarkers for chronic pain conditions and showed that their predictive efficacy heavily depended on psychological and social influences. These findings underscore the necessity of adopting a holistic approach in the development of biomarkers to enhance their clinical utility.

---

Chronic pain is a prevalent and complex condition that is difficult to measure due to its subjective nature and substantial variability between individuals and contexts ^1^. Consequently, developing biomarkers as objective measures of chronic pain has emerged as a high priority, given their potential to improve both the prognosis and management of pain ^2^. For instance, biomarkers could detect pain in vulnerable populations, stratify patients into homogeneous groups to improve treatment efficacy, or provide an objective assessment of interventions in randomized controlled trials. Biomarkers could also help define the pathogenesis of chronic pain conditions and identify novel targets for the development of new treatments ^3^. Despite their tremendous value, progress in identifying reliable biomarkers for chronic pain has been slow, and further research is needed to demonstrate their utility as diagnostic and prognostic tools.

Identifying biomarkers for chronic pain has been challenging, in part, because the severity of disease or injury is not a reliable indicator of the prognosis of pain. For instance, radiographic measures of joint degeneration in osteoarthritis fail to strongly predict the pain experienced by the patient ^4^. Other limiting factors hindering the identification of reliable biomarkers for pain include small sample sizes ^5^, lack of dataset generalizability ^6^, restricted availability of biological features, and the investigation of a limited range of pain phenotypes ^7^. In addition, the distinction between acute and chronic phases (*i.e.*, duration > 3 months) ^8^, its etiology (nociplastic, neuropathic, or nociceptive) ^9^, and its spatial extent (*i.e.,* localized or widespread) ^10^ further complicate the identification of comprehensive signatures for chronic pain.

Adding to the challenge, efforts toward biomarker identification have largely occurred in isolation, with biological predictors examined independently of other individual factors or environmental contexts. The interplay between biomarkers from different body systems and psychosocial factors has been largely ignored. To fully leverage biomarkers in the context of personalised medicine, a holistic understanding of pain through a multifactorial approach is necessary ^1^. The emergence of large prospective biobanks, such as the UK Biobank, provides a unique opportunity to concurrently evaluate numerous biomarkers and integrate them within a comprehensive framework for the classification and prediction of chronic pain.

In this study, we applied machine learning to multi-dimensional biological modalities including brain imaging, bone imaging, blood immunoassays, and genetics to derive candidate biomarkers for chronic pain. The relevant biomarkers were then contextualized by comparing their performance with psychosocial predictors of chronic pain that significantly contribute to the development of chronic pain ^10^. The synergy between biomarkers and psychosocial drivers of pain conditions was subsequently demonstrated to uncover the intricate biopsychosocial interactions that predict pain.

## Results

Our study utilized data from the UK Biobank to train and validate biomarkers for chronic pain related medical conditions and self-reported pain. The generalizability of the identified biomarkers was then tested using the All of Us Research Program (n = 27,151) and the Open Pain data repository (n = 250). A total of 493,211 participants were enrolled in the UK Biobank and completed the initial baseline visit. During that visit, participants were asked if they experienced pain that interfered with their usual activities in the last month at various body sites, including the head, face, neck/shoulder, stomach/abdominal, back, hip, knee, and pain all over their body. Participants who reported pain were then queried about pain lasting for more or less than 3 months (*i.e.*, chronic or acute). Participants were also asked about their medical conditions diagnosed from a physician and their use of medications. Blood samples were collected from participants to conduct whole genome sequencing and bioassay measurements. A subsample of 19,360 participants underwent a first follow up visit about four years after baseline where blood was recollected and a subsample of 48,079 participants underwent another follow-up visit about nine years after baseline, where brain imaging (magnetic resonance imaging; MRI) and bone imaging (dual-energy x-ray absorptiometry; DXA) were acquired. The timeline of data acquisition (T0: baseline, T1: about four years later, T2: about nine years later) and the sample sizes available are summarized in **Extended Data Fig. 1**. A schematic of the study aims is presented in **Fig. 1a**.

**Figure 1:**
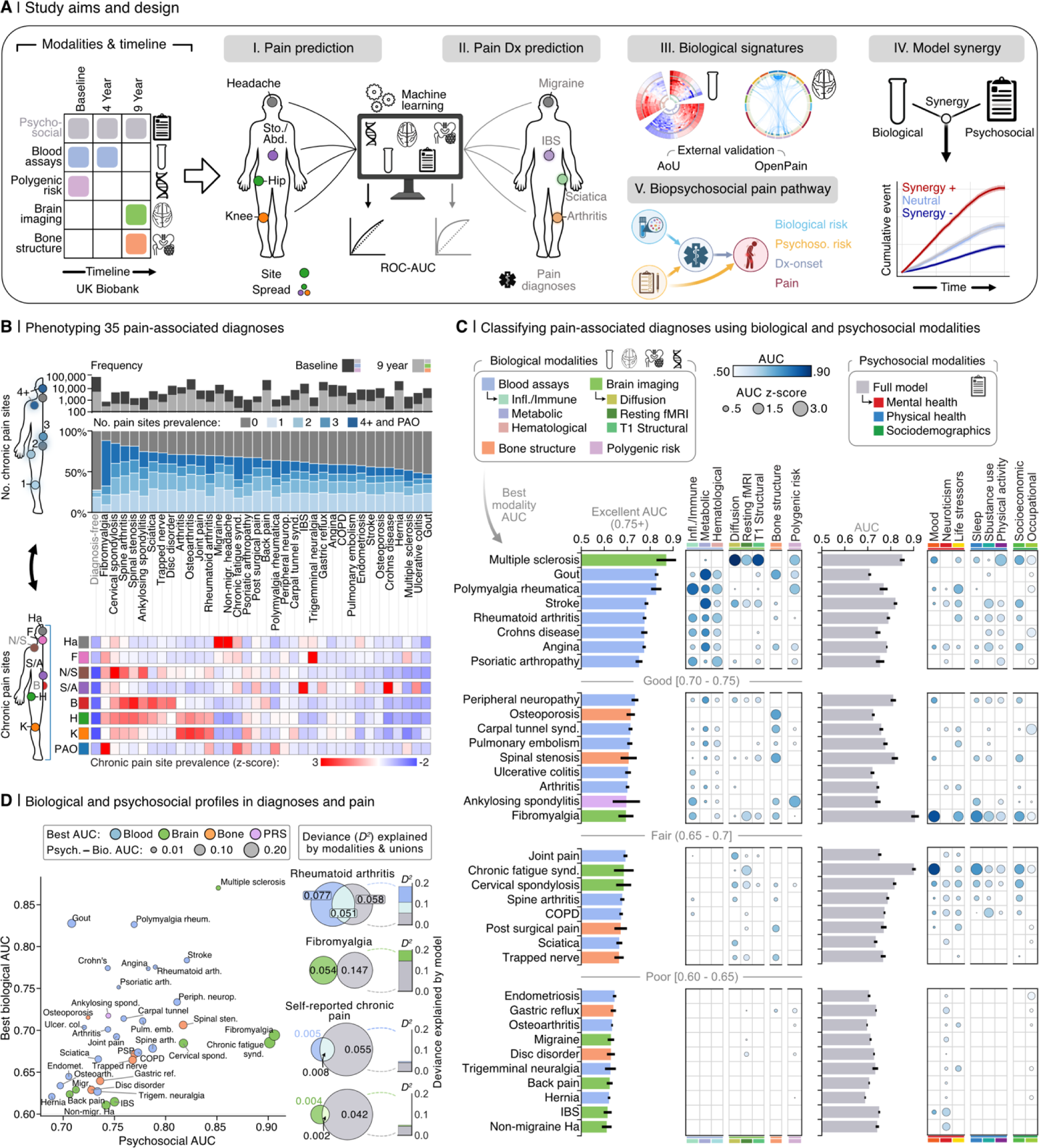
Classifying pain-associated diagnoses using biological and psychosocial modalities. **A.** Schematic illustrating the study workflow. **B.** Top: Barplots presenting the prevalence of chronic pain across 35 pain-associated diagnoses, categorized by the number of self-reported pain sites, and ordered by overall chronic pain prevalence. Above: Counts of diagnoses at baseline (black) and 9-year follow-up (grey). Bottom: A heatmap displaying the prevalence of pain sites for each diagnosis, normalized (z-score) across conditions for each specific site. The diagnosis-free control group is labeled in light grey for both plots. **C.** Barplots display the performance of models in classifying pain diagnoses in the test set, with error bars indicating the 95% confidence interval estimated from 1,000 bootstrap samples over 5 iterations of 5-fold Cross-Validation (CV). The bars represent the highest ROC-AUC scores achieved using biological (left) and psychosocial (right) modalities. The colors of the bars show the modality obtaining the highest AUC. Bubble heatmaps show ROC-AUC scores for modality subcategories, where applicable; bubble color indicates the absolute AUC score, and size reflects the z-score of the AUC relative to other diagnoses within a given modality or subcategory. Only diagnoses with z-scores above zero, indicating performance above the group mean AUC, are shown. For clearer visualization, diagnoses are grouped by their best biological modality performance (i.e. highest AUC score) into four categories, from Poor [0.60 - 0.65 AUC) to Excellent (0.75+ AUC). **D.** The scatterplot shows the comparison of AUC scores between the best biological and psychosocial modalities for each pain diagnosis, with points colored according to the best biological modality and labeled by diagnosis. Point size reflects the absolute AUC score difference, highlighting the discrimination discrepancy between biological and psychosocial factors for each diagnosis. Adjacent Venn diagrams depict unique and shared deviance explained (D2 ) in example pain diagnosis prediction (e.g., Rheumatoid arthritis & Fibromyalgia) and self-reported chronic pain prediction by the best biological (left circle) and psychosocial modality (right circle), with overlaps shown in the center. Corresponding stacked barplots convey the same information, with each segment color-coded by modality. Ha, headache; F, facial; N/S, neck or shoulder; S/A, stomach or abdominal; B, back; Hp, hip; K, knee; PAO, pain all over; COPD, chronic obstructive pulmonary disease, IBS, irritable bowel syndrome; PRS, polygenic risk score.

### Biomarkers for pain-related medical conditions

We initially aimed to identify biological signatures predictive of various medical conditions associated with chronic pain. Models underwent training through nested cross-validation, using logistic regression to classify individuals reporting pain versus those reporting no pain (**Extended Data Fig. 1**). These linear classifiers were trained on measures of inflammatory/immune, metabolic, and hematological assays to derive the blood biomarkers, measures from high-resolution T1-weighted imaging (grey matter), diffusion-weighted imaging (white matter), and resting-state imaging (functional connectivity) to derive the brain biomarkers, and measures from bone mineral content, shape, and density to derive the bone biomarkers. The genetic biomarkers were polygenic risk scores (PRS) derived at different thresholds from whole genome sequencing data (biological features are described in **Supplementary Tables 1-6**).

We selected 35 medical conditions in which the prevalence of chronic pain ranged between 48-80% of cases, most commonly presenting as multi-site pain (**Fig. 1a**). By comparison, the prevalence of chronic pain in participants without any diagnosed conditions was approximately 25% and typically localized to a single site. Medical conditions with pronounced multi-site pain exhibited symptoms in the neck, back, hip, and knee, whereas conditions where pain was concentrated around a single epicenter were localized in the head, face, and abdomen (**Fig. 1a**). Moreover, conditions such as fibromyalgia, chronic fatigue syndrome, and polymyalgia rheumatica were characterized by pain experienced all over the body (**Fig. 1a**). The demographics varied between pain-associated medical conditions and are shown in **Extended Data Fig. 2.**

The best predictions were obtained for conditions with a well characterized pathogenesis, such as multiple sclerosis (brain, AUC = 0.87), gout (blood, AUC = 0.83). and polymyalgia rheumatica (blood, AUC = 0.82). About half of the selected medical conditions could be predicted with good accuracy (AUCs > 0.70), including fibromyalgia (brain, AUC = 0.70), osteoporosis, spinal stenosis (bone, AUCs = 0.72, 0.70), peripheral neuropathy, and arthritis (blood, AUCs = 0.73, 0.70). The full weighted signatures for each of these biomarkers are presented in **Supplementary Fig. 1-10**. Overall, conditions with well-defined pathophysiology (e.g., gout, polymyalgia rheumatica) showed higher accuracy compared to those primarily defined by symptomatology (e.g., non-migraine headache, irritable bowel syndrome, back pain, **Fig. 1b**).

To better understand the pathogenesis of the various medical conditions, the performance of biomarkers (measured using their AUCs) within each biological category were normalized across medical conditions, after breaking down brain imaging and biochemical assays into their subcategories (**Fig. 1b**). Interestingly, the condition that was best predicted from polygenic risk scores was ankylosing spondylitis, a condition with well-established genetic contributions ^11^. Moreover, brain functional connectivity best predicted nociplastic pain conditions, such as fibromyalgia and chronic fatigue syndrome, which are hypothesized to be caused by central amplification of the nervous system ^12^. The comparison between biomarker subcategories also revealed that certain medical conditions could be independently predicted from multiple biological modalities (*e.g.*, ankylosing spondylitis, fibromyalgia, and polymyalgia rheumatica) while others were distinctly predicted from a single biological modality (e.g., chronic fatigue syndrome, osteoporosis, and ulcerative colitis). Sex differences in the performance of biomarkers across all modalities and medical conditions are detailed in **Extended Data Fig. 3**. Generally, differences in biomarker performance between sexes were minimal. Notable exceptions, however, included the bone-based marker for osteoporosis, which performed better in men, and the brain-based marker for peripheral neuropathy, which was more effective in women.

We next entered psychosocial features into the same machine learning analytic pipeline to classify each pain-associated medical condition. Most pain conditions were classified with fair to excellent accuracy, with conditions recognized as nociplastic being the most accurately classified, underscoring the significance of psychosocial factors in conditions like fibromyalgia and chronic fatigue syndrome (**Fig. 1b**). A closer examination of these psychosocial contributors, which could serve as either predictors or outcomes of the conditions, revealed mood and sleep disturbances as primary features for fibromyalgia, lower substance use as a notable characteristic of COPD, reduced physical activity as a marker of multiple sclerosis, and occupational factors as significant indicators of carpal tunnel syndrome. Using support vector machines and gradient boosting decision trees as alternative machine learning algorithms showed consistent results, although decision trees artificially improved the brain and blood-based biomarkers (**Extended Data Fig. 4**). Altogether, our results showed that a mosaic of biological and psychosocial markers coalesce in the manifestation of various conditions.

The summary plot presented in **Fig. 1c** shows the simultaneous performance of models trained on either biological or psychosocial features for each condition. These findings underscore the variance in predictive capability between biological and psychosocial factors across different conditions, enabling the identification of conditions predominantly predicted by biological markers (e.g., gout), those primarily predicted by psychosocial markers (e.g., non-migraine headache), and those where both types of markers play a significant role (e.g., polymyalgia rheumatica, rheumatoid arthritis, and stroke). The deviance explained – reflecting the reduction in discrepancy between the model’s predictions and the observed data – greatly varied depending on the medical condition. For instance, rheumatoid arthritis was most strongly determined from blood assays with a strong overlap with psychosocial factors whereas fibromyalgia was most strongly determined from psychosocial factors with little overlap with the brain biomarker (**Fig. 1c**). In the case of self-reported pain, the deviance explained was low and almost exclusively from psychosocial factors, emphasizing the limits of biological predictors for the subjective experience of pain.

### A validated blood-based signature for various chronic pain conditions

Biomarkers developed from blood immunoassays outperformed other categories of biomarkers, successfully predicting 13 pain-associated medical conditions with good accuracy (AUC > 0.70; **Fig. 1b**). Notably, these biomarkers demonstrated robust predictive performance on other conditions they were not originally trained to predict (**Extended Data Fig. 5**). We therefore trained a composite signature to concurrently predict these 13 medical conditions (**Fig. 2a**). The composite signature was comprised of strong positive coefficients (elevated in disease) for C-reactive protein, neutrophils, and gamma glutamyl-transferase, alongside notable negative coefficients (reduced in disease) for lymphocyte percentage, HDL cholesterol, and albumin (**Fig. 2b**). The effect of different medication categories (e.g., glucocorticoids, analgesics, statins) on the composite signature was tested by re-assessing the model’s performance after removing individuals who were taking medications from a specific category (**Extended Data Fig. 6**). The performance was generally stable, with the sharpest decreased in AUC observed upon excluding users of statins, analgesics, or antihypertensives (AUCs -0.05, -0.06, -0.06 respectively; **Extended Data Fig. 6**).

**Figure 2:**
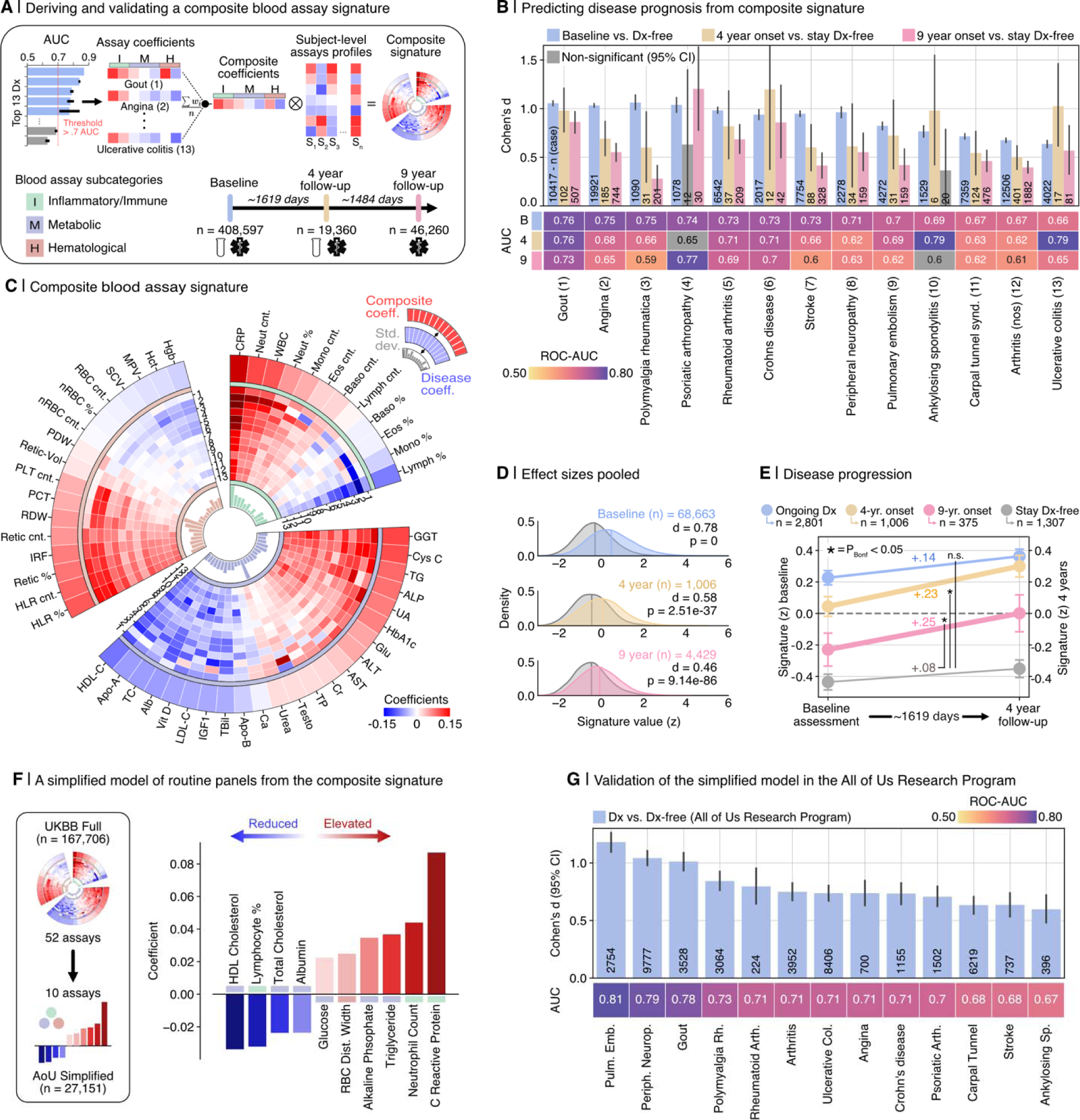
Deriving and validating a composite blood assay signature of pain-associated conditions. **A.** Above: Schematic illustrating the development of a composite blood assay-based signature derived from structure coefficients associated with 13 pain-related diagnoses. Below: Timeline and sample sizes from UK Biobank (UKB) data points utilized in the signature assessment. **B.** The composite signature’s diagnostic and prognostic efficacy for predicting 13 diagnoses is depicted using Cohen’s d and ROC-AUC, comparing diagnosed individuals to those diagnosis-free and individuals who develop a diagnosis to those staying diagnosis-free. Diagnostic accuracy is determined at the signature measurement time (baseline visit), while prognostic evaluations are conducted for individuals developing diagnoses within 4 and 9 years post-signature measurement. Error bars represent the 95% confidence interval estimated from 1,000 bootstrap samples. **C.** Circular graph depicting the composite signature’s structure. The outermost layer shows a heatmap of blood marker coefficients composing the composite signature. Middle layers present individual diagnosis marker coefficients, sorted by performance and numbered as in B. Inner layers display the standard deviation of marker coefficients across diagnoses. The graph is divided into three sections representing the blood assay subcategories: inflammatory/immune, metabolic, and hematological. **D.** Pooled effect sizes from C, measured by Cohen’s d, are calculated by comparing subjects diagnosed with any of the 13 diagnoses at baseline or developing a diagnosis at 4 or 9 years, against subjects who are diagnosis-free at baseline or remain diagnosis-free, respectively. **E.** The temporal changes in the predictive signature is quantified by comparing baseline and 4-year follow-up values among participants with a persistent diagnosis from baseline (Ongoing Dx), those who develop a diagnosis by the 4-year follow-up (4-year onset), those who develop a diagnosis by the 9-year follow-up (9-year onset), and those remaining diagnosis-free at the 4-year mark (Stay Dx-free). The significance of signature change between each group and the Stay Dx-free control group were estimated using linear mixed effects models, with adjustments for multiple comparisons using Bonferroni correction (*P*_bonf._ < 0.05). Error bars represent the 95% confidence interval, estimated from 1,000 bootstrap samples. **F,** A simplified model featuring the top 10 assays, selected based on their absolute structure coefficients from the composite signature, was developed and validated using data from the All of Us Research Program. **G,** The discriminatory power of the simplified model was quantified using Cohen’s d and ROC-AUC values, with error bars indicating the 95% confidence interval calculated from 1,000 bootstrap samples. Inflammatory/Immune markers: CRP, C-reactive protein; Neut, neutrophil; WBC, white blood cell; Mono, monocyte; Eos, eosinophil; baso, Basophil; Lymph, lymphocyte Metabolic markers: GGT, gamma glutamyl transferase; Cys C, cystatin C; TG, triglyceride; ALP, alkaline phosphatase; UA, uric acid; HbA1c, glycated hemoglobin, Glu, glucose; ALT, alanine aminotransferase; AST, aspartate aminotransferase; Cr, creatinine; TP, total protein; Testo, testosterone; Ca, Calcium; ApoB, apolipoprotein b; TBil, total bilirubin; IGF1, insulin-like growth factor; LDL-C, low density lipoprotein cholesterol; Alb, Albumin; TC, total cholesterol Hematological markers: HLR, high light scatter reticulocyte percentage; Retic, reticulocyte; IRF, immature reticulocyte fraction; RDW, red blood cell distribution width; PCT, platelet count; PLT, platelet count; PDW, platelet distribution width; nRBC; nucleated red blood cell; SCV, sphered cell volume; MPV, mean platelet volume; Hct, hematocrit; Hgb, hemoglobin; Dx, diagnosis; UKBB, UK Biobank; AoU, All of Us Research Program.

Using longitudinal data from follow-up visits at 4 (T1) and 9 (T2) years, the composite signature measured at baseline (T0) predicted newly developed diagnoses at T1 (average AUC 0.70; range: 0.62-0.79), with robust predictions for over half of the conditions (AUCs > 0.70). The composite signature measured at T0 could also predict newly diagnosed medical conditions at T2 that were not yet diagnosed at T1 (*e.g.*, Crohn’s disease, gout, psoriatic arthropathy AUCs > 0.70; **Fig. 2c**). Effect sizes are shown in **Fig. 2d** and longitudinal changes in the expression of the composite signature are shown in **Fig. 2e**. A simplified version of the composite signature (**Supplementary Table 1**) was validated in the All of Us Research Program (cross sectional; **Fig. 2f**), obtaining similar performances and demonstrating its generalizability across conditions in a cohort from a different country. Notably, participants in this validation cohort were younger (**Extended Data Fig. 7**) and therefore less likely to use statin or antihypertensive medication prescriptions.

### A validated brain-based signature for nociplastic conditions

In contrast to blood-based biomarkers, biomarkers based on functional connectivity were largely specific to distinctly nociplastic pain conditions such as fibromyalgia, chronic fatigue syndrome, and widespread pain (AUCs = 0.64-0.66). The weights of the logistic regression models for these three multivariate signatures are presented in the **Supplementary Fig. 5**. The encoding maps showing the univariate structure coefficients of nodes associated with the model prediction are presented in **Fig. 3b**.

**Figure 3:**
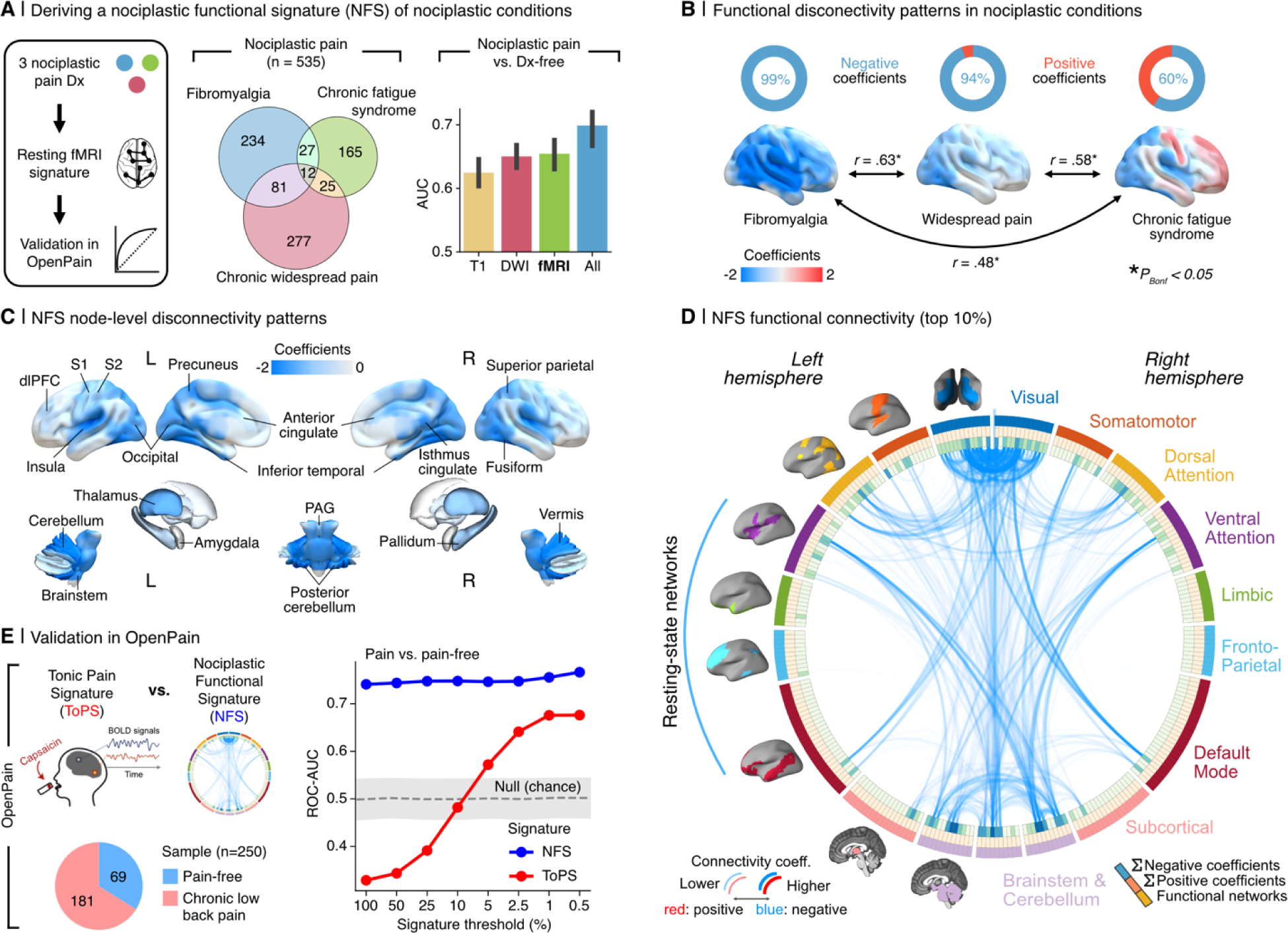
Deriving and validating a multivariate functional connectivity signature of nociplastic pain. **A.** Schematic describing the steps implemented to develop the nociplastic functional signature (NFS). Venn diagram shows the sample sizes and sample overlap among the three nociplastic conditions—Fibromyalgia, Chronic Fatigue Syndrome, and Chronic Widespread Pain—used to derive a combined nociplastic pain phenotype (n = 535). Barplots show ROC-AUC results from models trained on brain imaging modalities to classify this phenotype, with error bars denoting the 95% confidence intervals from 1,000 bootstrap samples over 5 iterations of 5-fold cross validation. **B.** Top: Donut plots display the proportions of positive versus negative structure coefficients from models predicting the component nociplastic conditions, based on resting-state functional Magnetic Resonance Imaging (rsfMRI) data. Bottom: Cortical surface renderings visualize connectivity, thresholded to highlight the top 25% of structure coefficients, which represent the sum of dynamic conditional correlation across brain parcels. Arrows interlinking the cortical renderings depict the association (two-sided Pearson correlation, all P < 0.05 Bonferroni corrected) between the complete unthresholded vectors of structure coefficients (parcel to parcel connectivity) for the respective conditions. **C.** Visualization of nociplastic signature connectivity, thresholded to emphasize the top 25% of structure coefficients that denote the sum of dynamic conditional correlations across brain parcels. **D.** Circos plot illustrating the top 10% of connectivity links (represented by structure coefficients) that constitute the nociplastic signature from the resting-state fMRI model, mapped across the canonical resting-state networks. **E.** Validation of the resting-state fMRI nociplastic signature (NFS) in four aggregated external cohorts from the OpenPain repository (n = 250), benchmarked against a pre-validated neural signature of capsaicin-induced sustained pain (Tonic Pain Signature, ToPS). Performance in discriminating pain vs pain-free groups across various densities of NFS and ToPS within the OpenPain cohorts is depicted in the plot to the right along with the standard deviation band of a null classification model generated from 10,000 permutations of the NFS. Nocip, Nociplastic; T1, T1 structural brain imaging; DWI, Diffusion Weighted Imaging; fMRI, function Magnetic Resonance Imaging; All combines the features from the T1, DWI, and fMRI brain imaging modalities; dlPFC, dorso-lateral prefrontal cortex; S1, primary somatosensory cortex; S2, secondary somatosensory cortex.

A new brain-based model was developed to predict a composite phenotype that encompassed all three nociplastic conditions. The weights of the signature are presented in **Supplementary Fig. 7**, and the structure coefficients of edges associated with the model prediction are depicted in a circular plot in **Fig. 3d** and brain rendering in **Fig. 3c**. The conditions were characterized by a general pattern of dysconnectivity between brain regions, notably involving the visual cortex, the brainstem and cerebellum, the dorsal and ventral attention networks, and the sensorimotor network. This brain-based signature, termed nociplastic functional signature (NFS), was validated in four distinct external datasets available on the Open Pain repository, which included individuals with chronic pain who predominately reported high levels of pain severity and impact (**Extended Data Fig. 7**). We then compared the performance of our brain signature with the Tonic Pain Signature (ToPS), an existing brain-based signature trained on capsaicin-induced pain that effectively classified clinical pain in cohorts from the Open Pain repository ^6^. The signatures were thresholded across different densities (100% - 0.5%) and then applied in the Open Pain repository to classify individuals with chronic pain from pain-free individuals (**Fig. 3e**). While both NFS and the ToPS obtained fair to good performances when restricted to the most important functional connections, the NFS was stable across all thresholds.

### Biomarkers for self-report pain

We have identified various biomarkers effective in classifying chronic pain associated medical conditions. Our findings however suggest that biomarkers’ performance may be more limited for self-reported pain. We therefore trained new models to classify the subjective report of presence or absence of pain, regardless of its location on the body. Here, biomarkers from either brain, bones, blood, or genes all demonstrated weak ability to distinguish between participants with and without pain, indicating their unreliability in pain prediction (chronic pain Vs. pain free: all AUC between 0.55-0.59; acute pain Vs pain free: all AUC between 0.52-0.54; **Fig. 4c**). Training separate models to predict pain located at each body site showed minimal improvement in performance (all AUC < 0.62; **Fig. 4d**). However, when models were trained to predict pain reported “all-over-the-body”, all biological categories, with the exception of genetic PRS, showed increased performance (AUC = 0.66-0.69). This contrasts with models trained on psychosocial factors including mood, sleep, life stressors, neuroticism, substance use, physical activity, and socioeconomic factors (psychosocial features are described in **Supplementary Table 6**), which performed well across all body sites (AUC > 0.70, **Fig. 4d**) and exhibited excellent accuracy for pain reported all-over-the-body (AUC = 0.92). Using alternative machine learning algorithms did not improve the prediction of self-reported pain (**Extended Data Fig. 4**). Our findings show that candidate biological markers did not predict chronic pain effectively unless pain was spread across body sites. Conversely, psychosocial factors offered a more reliable prediction of self-reported pain.

**Figure 4:**
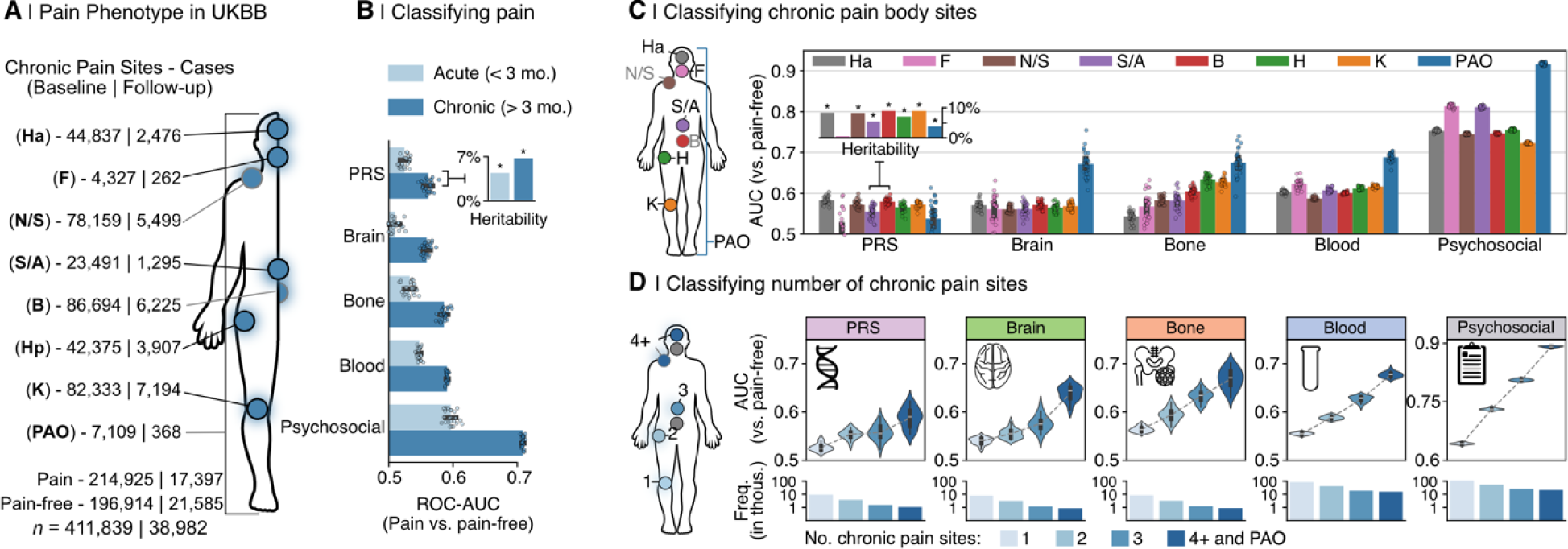
Classifying chronic pain phenotypes using biological and psychosocial modalities. **B.** Anatomical body map of chronic pain sites and their counts for the full (baseline) sample and for individuals with a follow-up visit 9 years later. **C.** Performance of machine learning models in classifying participants reporting acute (light blue) and chronic (dark blue) pain from pain-free in left out test sets (n_chronic_□=□2,679-42,985; n_acute_□1,012-16,259), shown via Area under the curve of the receiver operating characteristic (ROC-AUC) scores. Error bars show 95% CI from 1,000 bootstrap samples of 5 iterations of 5-fold cross-validation (25 total AUC scores). Also included are heritability estimates from Genome Wide Association Studies (GWAS) for both acute and chronic pain types. **D.** Left: Body map of chronic pain sites. Right: Barplots grouped by modality, showing machine learning model performance in distinguishing subjects with specific pain sites from pain-free controls in the same manner as C. **E.** Left: Body map showing aggregation of chronic pain sites to depict a phenotype based on the number of self-reported pain sites (i.e., anatomical pain spread), ranging from 1 to 4 or more distinct sites. Right: Violin plots by modality show test set performance (ROC-AUC) of models classifying chronic pain spread versus pain-free. Beneath: Barplots indicate the frequency of each pain spread phenotype in modality-specific models. IBS, irritable bowel syndrome; Dx, diagnosis; UKBB, UK Biobank; Ha, headache; F, facial; N/S, neck or shoulder; S/A, stomach or abdominal; B, back; Hp, hip; K, knee; PAO, pain all over; PRS, polygenic risk score; * denotes a Bonferroni-corrected P-value of less than 0.05.

We then trained separate biomarkers to predict the number of pain sites reported by the participant, varying the extent of pain spreading as the target instead of the anatomical location (**Fig. 4e**). Here, each biomarker showed a consistent increase in its performance as the number of pain sites was increased (average improvement in AUCs between single site and 4+ sites = 0.10; **Fig. 4d**). Feature encoding maps of biomarkers trained on varying number of pain sites were generally consistent, suggesting that pain spreading didn’t modify the biological signature but instead amplified its expression (**Extended Data Fig. 8**). Our results suggest that biomarkers showed increased sensitivity to the spreading of pain rather than to its specific location, although the performance remained relatively poor compared to psychosocial factors.

### Synergy between biological and psychosocial markers for pain

Lastly, we aimed to generate a holistic biopsychosocial framework by examining the interactions between biomarkers and psychosocial factors in relation to the onset and progression of chronic pain (**Fig. 5a**). To investigate this interaction, participants were grouped into five quintiles representing the expression of their pooled biomarker and their psychosocial risk. We extracted log probabilities from our top-performing models as individual risk scores, reflecting the likelihood of subjects having certain conditions. These scores were then averaged to generate a pooled risk score for each subject, effectively summarizing their overall biomarker and psychosocial risk profile. **Fig. 5b** shows the expression of the pooled risk scores and the odds ratio of diagnosis for the 13 medical conditions predicted from that risk score, as initially presented in **Fig. 3b**. Being in the highest quintile for either biomarkers (H) or psychosocial factors (H) elevated the risk of having the medical condition, as indicated by the odds ratio (OR) range between 6 and 18. However, being in the highest quintile for both (H-H) dramatically amplified the risk, with the range of odds ratios escalating between 18 and 42, as shown using a log scale in **Fig. 5c**. This effect was also observed in cross sectional data when varying the biomarker modality and using the medical conditions best predicted from those biomarkers: using the brain-based risk common to nociplastic conditions for fibromyalgia and chronic fatigue syndrome (**Fig. 5d**) or bone-based risk for osteoporosis, spinal stenosis, carpal tunnel syndrome, and gout (**Fig. 5e**). Overall, the synergy between biological and psychosocial markers was consistently observed across different biomarker modalities and pain-associated medical conditions, significantly enhancing the prediction of various medical conditions.

**Figure 5:**
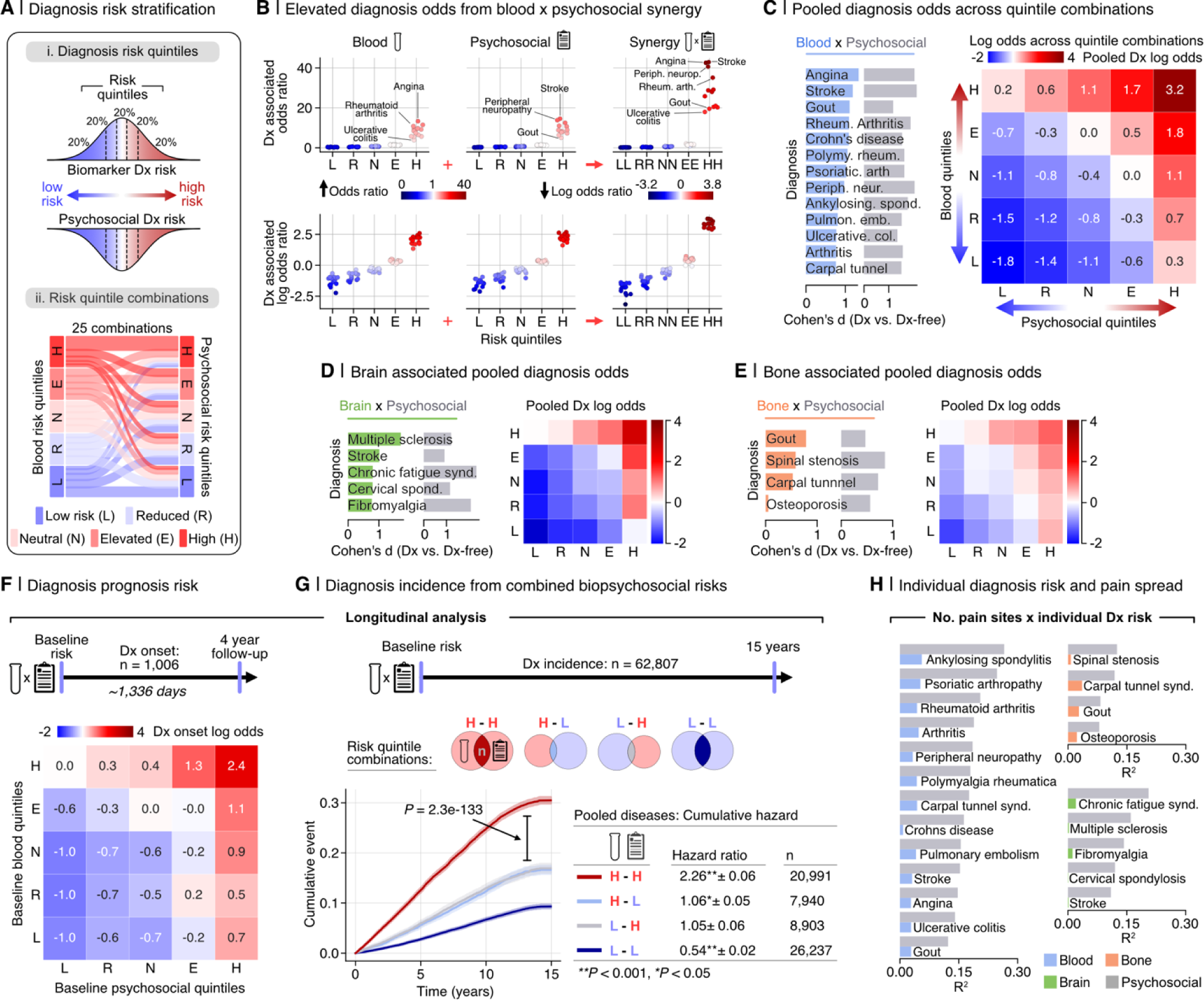
Assessing biopsychosocial synergy in the prognosis of pain-associated medical conditions. **A.** Overview of the development of diagnosis risk stratification models based on pooled probabilities from biological (blood, brain, bone) and psychosocial modalities. (i) Biological and psychosocial risk scores were segmented into quantiles for stratification. (ii) A Sankey diagram visualizes the possible combinations blood and psychosocial risk quintiles; this operation is similarly conducted for bone and brain risk models. **B,** Top: Diagnosis-associated odds ratios for each diagnosis are computed for participants within each risk quantile of blood assay risk scores, psychosocial risk scores, and combined risk scores, in comparison to all other participants. Bottom: The odds ratios are transformed to log-odds to elucidate the protective effect associated with lower risk quantiles. **C,** The performance of the pooled risk scores for each diagnosis was measured against diagnosis-free participants using Cohen’s d effect size. The heatmap displays the log-odds ratios of having a diagnosis across all risk quintile combinations, highlighting the synergistic impact of high combined biological and psychosocial risks and the protective effects conferred by lower combined risks. The analysis performed in C was replicated for diagnoses most accurately classified by the brain (**D**) and bone (**E**) modalities. **F,** Log-odds ratios depict the synergy between blood assay biomarkers and psychosocial factors in disease prognosis 4 years later. **G,** Kaplan-Meier curves show the cumulative incidence of receiving a diagnosis up to 15 years following baseline, segregated into four groups according to combinations of blood and psychosocial risk quantiles (High-High, High-Low, Low-High, and Low-Low). Hazard ratios are calculated using Cox proportional hazard models, while the P value of the differences between groups is calculated using a two-sided log-rank test. **H,** Barplots plots illustrate the association between biological and psychosocial risk scores for blood (left), bone (top right), and brain (bottom right) risks with categories of pain site spread (ranging from 1 to 4+ sites) The R² values indicate the strength of these associations, as determined by Spearman’s rank correlation; (* indicates p < 0.001).

Concordant results were obtained using the blood risk score in the longitudinal data (**Fig. 5f)**, as new diagnoses over a period of 15 years occurred more rapidly in the H-H group (Hazard Ratio (HR): 2.26 (2.20-2.32) compared to participants in either H-L (HR: 1.06 (1.01-1.11)) or L-H (HR: 1.05 (0.99-1.11)) or L-L groups (HR: 0.54 (0.51-0.56); **Fig. 5g**). Thus, participants with high biomarker and psychosocial risk (H-H quintiles at baseline) exhibited a significantly increased incidence of a new medical condition compared to subjects with high biomarker but low psychosocial risk scores (H-L quintiles at baseline). These findings imply the same biomarker performs differently depending on the psychosocial context in which it is being tested.

One of the important limitations of these biomarkers was their inability to explain the subjective experience of pain associated with the medical condition (**Fig. 5h**). For each medical condition, the variance in the number of pain sites was better explained from the psychosocial risk score (r^2^ ranging from 0.07-0.28) than the expression of the biomarker (best modality: r^2^ ranging from 0.00-0.06). These findings were consistent across all categories of biomarkers and medical conditions evaluated. We conclude that biomarkers and psychosocial factors do not act in isolation but synergize to influence the development and prognosis of pain-related medical conditions. However, solely psychosocial factors reliably determine the presence, the bodily distribution, and impact of chronic pain.

The blueprint of this holistic framework for the prediction of chronic pain is illustrated by applying structural equation modeling to longitudinal data available in the UKBB (**Fig. 6)**. Here, the blood-based risk score, the psychosocial risk score, and their interaction measured at baseline uniquely contributed to the development of rheumatoid arthritis (RA) over a period of 10 years. Pain outcomes were then extracted from the Online pain questionnaire collected in 2019. The model shows that biomarkers, psychosocial risk factors, and their combined effects explained unique variance in the development of RA. However, only the RA diagnosis and the baseline-measured psychosocial risk factors were directly linked to pain intensity, spread, and disability.

**Figure 6:**
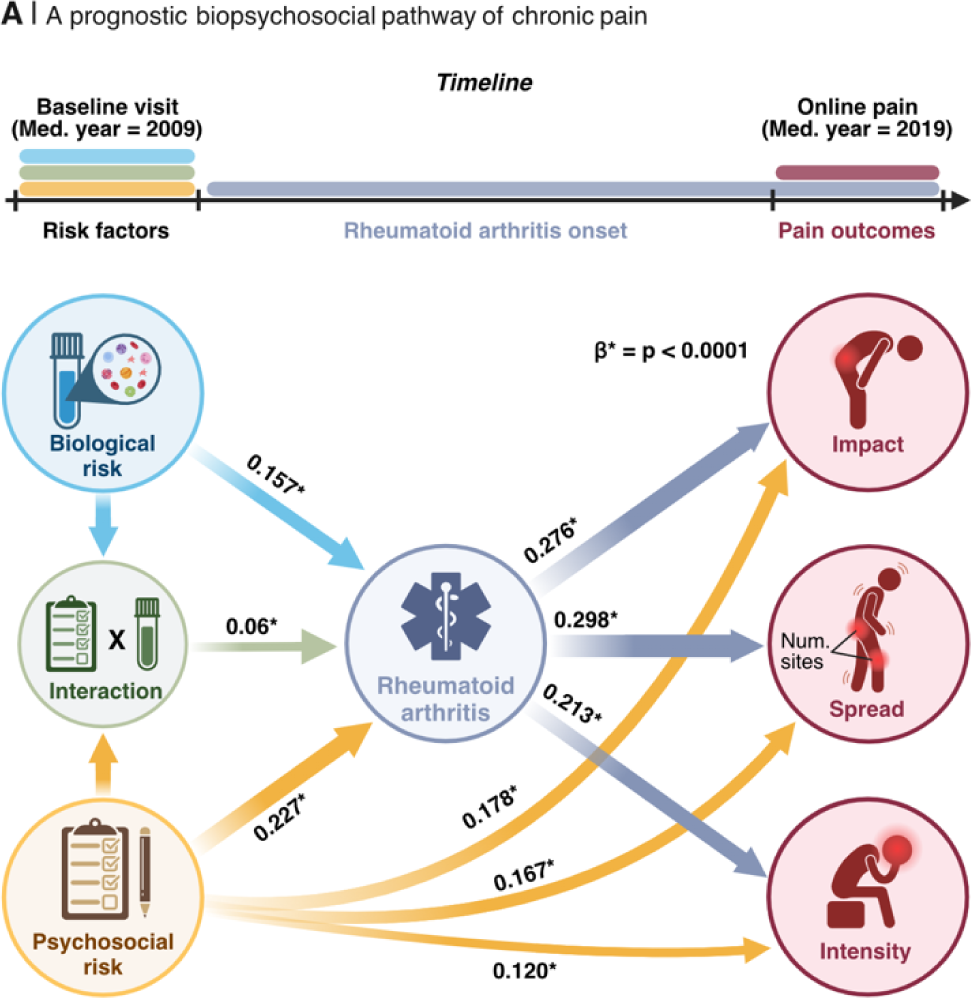
A holistic biopsychosocial framework for the development of chronic pain. A framework for the development of chronic pain over time is depicted through a data-driven structural equation model utilizing longitudinal data. Baseline biological risk is calculated using a blood risk score for rheumatoid arthritis, derived from 52 blood markers encompassing inflammatory and immune, metabolic, and hematological assays. Baseline psychosocial risk is quantified using a psychosocial risk score for rheumatoid arthritis, derived from 90 pain-agnostic features that include mental, physical, and sociodemographic factors. Pain diagnosis denotes the onset of healthcare diagnosed rheumatoid arthritis between the initial risk assessment and the online follow-up pain questionnaire roughly 10 years later. Pain outcomes encompass the interference of chronic pain across several dimensions (pain impact), the count of self-reported chronic pain sites (pain spread), and the rating of the worst pain in the last 24 hours (pain intensity) at the online pain questionnaire. Arrows are labeled with regression coefficients derived from the structural equation modeling. Arrows are only drawn for significant associations. *P < 0.0001; Bootstrap (1,000 iterations) tests were used for significance testing of regression coefficients.

## Discussion

This study aims to identify and compare various biomarkers for chronic pain using biological measures from different body systems. Overall, we found that biomarkers for self-reported pain performed poorly, in contrast to those targeting the medical conditions underlying the pain that demonstrated strong diagnostic and prognostic potential. Importantly, biological markers and psychosocial factors synergize to significantly influence the prevalence of chronic pain conditions, evident in both cross-sectional and longitudinal assessments. This highlights the importance of adopting a holistic framework to identify biomarkers for chronic pain and contributes to refining the biopsychosocial model for the prediction of future chronic pain.

One of the main results of our study is that candidate biomarkers for self-reported pain performed poorly. This outcome seems contradictory to the extensive literature associating clinical pain with biological anomalies ^13–15^ but several factors can explain this discrepancy. First, our findings do not imply the absence of such associations; they instead show that these factors alone are inadequate for predicting chronic pain in new participants. Second, outside the realm of genetics, most previous studies comparing chronic pain patients to pain free controls made no distinction between the medical condition causing the pain and the subjective pain report. Although our results show that pain symptoms were most strongly predicted from psychosocial factors, they also show that the underlying condition often could be predicted from biomarkers. Finally, the generalizability of several studies that report the identification of biomarkers for chronic pain may be limited due to their small sample sizes, which can lead to overfitting and inflated effect sizes^16^. This is consistent with recent findings in the UKBiobank, where minor effect sizes (Cohen’s d < .15) were observed in grey matter reduction when large groups of chronic pain patients were compared to those without pain^17^. For these reasons, biological abnormalities documented in the pain literature remain in the realm of explanation rather than prediction ^18^. The novelty of our study lies in testing the predictive capacities of various biomarkers in left out participants. If successful, the identified biomarkers were then further evaluated in a new group of participants outside of the UK Biobank. This is especially important given that accurate predictions of clinical outcomes within trials in which models were developed often showed no better than chance level when tested in out-of-sample patients ^19^.

The poor performance of biomarkers for self-reported pain is likely to be explained by the heterogeneity of the conditions (e.g., ankylosing spondylitis, spine arthritis, or sciatica) causing pain at a common body site (e.g., back). Yet, pain and its spatial spread were still best predicted from psychosocial factors after selecting a single medical condition for which an effective biomarker was identified (e.g., rheumatoid arthritis, ankylosing spondylitis, Crohn’s disease). This highlights the challenges of identifying biomarkers for a subjective measure characterized by non-linearity between its biological manifestation and its subjective experience. Our results demonstrate that chronic pain cannot be fully accounted for from the pathogenesis of the medical condition, which aligns with abundant previous research and clinical experience ^20,21^. This phenomenon was consistent regardless of the body systems used to train the biomarkers or the etiology of the predicted medical conditions. These findings emphasize that overlooking the role of psychosocial factors would severely constrain our capacity to identify future biomarkers that would generalize across individuals. The limited ability of biological markers, such as brain imaging, for clinical predictions is not exclusive to the field of pain research. The field of psychiatry has encountered a comparable problem with depression ^22^ for which no clinically useful brain-based biomarkers have yet been identified ^23^.

Nonetheless, efforts in chronic pain research have historically aimed to identify biomarkers generalizable to clinical pain (i.e., presence of chronic pain) ^2,14,23^. Our findings suggest a more targeted approach is necessary, focusing on biomarkers specific to the medical conditions associated with clinical pain. Chronic pain originates from various mechanisms— nociplastic, neuropathic, inflammatory, and nociceptive—each characterized by unique pathological features embedded in different bodily systems ^9,24,25^. By concentrating on specific pain-related conditions, biomarkers can more accurately target these distinct mechanisms. For example, a biomarker developed for spinal stenosis can specifically target its neuropathic origins, unlike a biomarker for general back pain, which must account for a variety of underlying mechanisms. Another example is how inflammatory processes in rheumatoid arthritis and Crohn’s disease are reflected by blood biomarkers, which show strong positive coefficients for indicators such as C-reactive protein and leukocytes (**Supplementary Fig. 1**). Additionally, this method allows for the development of composite biomarkers suitable for use across multiple diagnoses that share similar mechanisms. By breaking down chronic pain into its clinical subtypes, we can identify and leverage mechanistic overlaps to craft biomarkers that span multiple conditions. We demonstrated this through a composite blood signature predicting several inflammatory, neuropathic, and nociceptive conditions (**Fig. 2**), as well as a functional connectivity signature for nociplastic pain (**Fig. 3**), demonstrating that despite condition-specific biological markers, a shared underlying pathology can offer sufficient predictive capability.

Lastly, anchoring biomarker discovery in diagnoses rather than subjective pain reports provides a more objective and consistent basis for biomarker research. The complexities surrounding the definition of chronic pain—its duration, characteristics, and subjective experience—presents challenges in biomarker validation ^26^. Focusing on clinically recognized pain diagnoses offers a more stable framework, sidestepping the variability of subjective pain reports with the more consistent symptomatology of diagnosed conditions. This shift towards clinical phenotypes in biomarker research promises to refine our understanding of the pathogenesis and treatment of chronic pain, moving beyond the limitations of subjective pain assessment.

Our results do not undermine the importance of subjective pain reports or their clinical relevance; patient-reported pain and psychosocial factors, such as mental and physical health, remain central targets for treatment. However, we found that even effective biomarkers for specific pain diagnoses, like gout and rheumatoid arthritis do not correlate directly with these subjective pain reports (**Fig. 5,6**). Instead, psychosocial factors emerge as key to understanding reports of pain severity, its impact, and spread (**Fig. 6**), highlighting the often-overlooked significance of psychosocial contexts in biomarker development for chronic pain.

Reliable biomarkers for pain-causing pathology were unavailable for most forms of pain conditions ^2^. Here, the most consistent predictions were obtained using blood immunoassays, where notably, higher C-reactive protein, neutrophils, and cystatin-C and lower lymphocyte percentage, HDL cholesterol, and albumin predicted various inflammatory, musculoskeletal, and neuropathic pain conditions—a conclusion supported by clinical and research evidence ^27–30^. The signature is therefore likely to reflect a systemic state of inflammation or general health deterioration that is unspecific to any individual pain condition but rather characterizes a broader state of pathology. There is precedent for such multidisease models, as evidenced by Buergel et al. (2022), who applied deep learning to metabolomics to assess the risk of various common diseases ^31^. Here, our study employed routine immunoassays—such as C-reactive protein for inflammation, uric acid for metabolic assessments, and neutrophil counts for immune response— targeting mechanisms central to pain-associated conditions. We then applied interpretable linear models and encoding maps to specifically trace their impacts on the conditions. Importantly, the composite signature trained to simultaneously predict multiple pain conditions performed well cross sectionally and longitudinally and generalized in an external large dataset, the All of Us Research Program. Our results also uncover brain-based markers that were more specific for pain conditions characterized by widespread pain, such as fibromyalgia, chronic fatigue syndrome, or pain experienced all over the body. These findings align with the literature suggesting that widespread pain observed in nociplastic conditions arises from altered brain function ^12,32^. The structure coefficients of the brain connectivity associated with the prediction of the model revealed lower functional connectivity between a distributed set of brain regions, a phenomenon that has previously been reported in fibromyalgia patients ^33^. The NFS was successfully validated in four external datasets available in the Open Pain repository.

The comprehensive set of biomarkers derived in this study to classify various medical conditions and pain reports highlights the importance of a holistic framework for the prediction of chronic pain. Our results show that biological and psychosocial risk factors additively influence the likelihood of receiving a medical diagnosis, indicating that changes in environmental, behavioral, and psychosocial factors either preceding or concurrent with disease onset, are crucial for accurately classifying chronic pain conditions. By combining a blood draw measuring the composite signature with a brief questionnaire predicting pain spread such as the risk Risk of Pain Spreading (ROPS)^10^, the holistic approach outlined in this study could be leveraged to significantly improve the prognosis of pain-associated conditions and self-reported pain symptoms like spread, severity, and interference.

Altogether, our results imply that painful medical conditions can be accurately predicted from the synergy between biological and psychosocial factors, but that pain report was solely predicted from psychosocial factors. Thus, our findings challenge the established idea of a reliable biological marker that could, on its own, detect or predict the subjective experience of chronic pain reported by patients. Instead, our research suggests that future biomarker discovery efforts in chronic pain should incorporate relevant psychosocial factors—such as occupational influences in carpal tunnel syndrome or lifestyle factors in Crohn’s disease—into their development protocols. This holistic approach aims to enhance the diagnostic and prognostic utility of biomarkers and support personalized treatment strategies. By integrating biological and psychosocial insights, this comprehensive strategy promises not only to refine diagnostic accuracy but also to identify treatments that address both the mechanistic underpinnings and the psychosocial dimensions of pain, ultimately reducing patient suffering.

## Online Methods

### Overview of the UK Biobank population

The UK Biobank project is a large-scale, prospective, and ongoing study initially established to allow extensive investigation on genetic factors and lifestyle determinants of a diverse range of common diseases of middle-aged and older adults^1^. To recruit the intended sample size of approximately 500,000 participants, over 9 million invitations were sent to individuals registered in the UK National Health Service (NHS) with an inclusion age range of 40-69 years old and based on their living within a reasonable distance from an assessment center. Baseline recruitment and data collection from 503,317 participants who consented to join the study took place between 2006 and 2010 in 22 assessment centers throughout Scotland, England, and Wales^2^. All participants gave written, informed consent, and the study was approved by the Research Ethics Committee (REC number 11/NW/0382). Further information on the consent procedure can be found elsewhere. Subsets of baseline participants were invited later for follow-up visits and/or were asked to provide data on certain online questionnaires at certain timepoints. The following datasets from different timepoints are used to address different aims of our study.

### Biological Modalities

Broad biological modalities indexing four diverse domains of physiological health were selected for analyses. Biological measures included blood immunoassays, polygenic risk scores, bone structure estimates derived from Dual-energy X-ray absorptiometry (DXA) scans, and brain imaging phenotypes. For certain analyses, we segmented blood and brain modalities into subcategories. Specifically, blood was divided into three distinct assay types: those assessing inflammatory or immune functions, metabolic functions, and hematological functions. Brain imaging was divided into: T1-weighted MRI for anatomical insights, diffusion MRI (dMRI) for white matter architecture, and resting-state functional MRI (fMRI) for brain connectivity.

The availability across study visits, derivation, and processing of each modality are detailed below:

### Blood immunoassays (52 features, N = 476,787)

The UK Biobank measured 31 haematological (https://biobank.ctsu.ox.ac.uk/crystal/crystal/docs/haematology.pdf) and 30 biochemical (https://biobank.ndph.ox.ac.uk/showcase/ukb/docs/serum_biochemistry.pdf) blood assays using high fidelity haematology, immunoassay, and clinical chemistry analyzers. For the purposes of this study, 52 distinct assays were selected measuring a range of physiological functions including liver health (Gamma-Glutamyl transferase), kidney function (Cystatin C), systemic inflammation (C-Reactive protein), and immune activation (Leukocyte count), among others. Blood assays were collected twice: initially during the baseline evaluation of 502,494 participants and subsequently in a subset of 20,193 participants who returned approximately 4 years later (range: 1-6 years), hereafter referred to as the 4-year follow-up. At both time points, participants missing data for 22 or more assays (constituting over half of the selected assays) were excluded from further analysis. For the remaining participants with any missing data (up to 13% of the total sample, or a maximum of 62,216 individuals), we imputed missing values using the median of the available data. This process yielded a final analytic cohort of 476,787 individuals at baseline and 19,360 at the 4-year follow-up. To address distributional biases in raw assay measures, we applied logarithmic transformation to all immunoassay data prior to model training.

### Polygenic Risk Scores (20 features, N = 408,597)

Blood samples collected at the baseline visit allowed different types of assays to be performed, including genetic data. A total of 488,118 underwent genotyping and have available phenotype information. Participants of similar genetic ancestry (Field: 22018), participants of non-European ancestry (Field: 22006), as well as participants who failed quality control (Field: 22010) were excluded from genetic analysis, resulting in 408,597 participants. This population was split into a training and a testing set, with the latter comprising individuals who attended the 9-year follow-up (n = 40,898), as conducted in Tanguay-Sabourin et al. 2023^6^. Consequently, polygenic risk scores were estimated within the training set of 367,699 participants and evaluated on the testing set of 40,898 participants as described below:

#### Genome Wide Association Studies (GWAS)

For each training set across each pain phenotype and pain-associated diagnosis, we conducted genome-wide association studies (GWAS) using regenie^26^. Regenie was selected for its ability to address cryptic relatedness among UK Biobank (UKB) participants and manage case/control imbalances. Covariates included sex, age, age squared, genotyping array, the first 40 genetic principal components, and dummy-coded recruitment sites. Sex was determined based on genetically ascertained sex (XY=man, XX=woman; field 22001). In regenie’s step 1, 93K SNPs identified by the UKB for kinship estimation were analyzed. For step 2 in regenie, approximately one million SNPs were evaluated, selected from the European-specific subset of the pan-UK Biobank project. These SNPs are high-quality HapMap 3 variants that meet five criteria: located in autosomes, outside the MHC region, bi-allelic, with INFO > 0.9, and MAF > 1% in both UKB and gnomAD datasets. Individuals retained for analysis were of European ancestry (field 22006), excluding any with failed genotyping quality, unusual heterozygosity, sex chromosome anomalies, or those who had withdrawn from the study by December 2021 (according to the sample-QC file from resource 531). Lastly, narrow-sense heritability estimates for each pain phenotype and diagnosis were derived using LDSC from the GWAS summary statistics^27^.

#### Polygenic Risk Scores (PRS)

Polygenic Risk Scores (PRS) were constructed ensuring that included SNPs were not in linkage disequilibrium, starting from the genotyping data in PLINK format provided by the UK Biobank. Utilizing PLINK,^28,29^ we removed all non-rsID SNPs as previously described, and excluded SNPs with MAF < 0.001, genotyping error rates > 1%, and Hardy-Weinberg equilibrium P-value < 10^-12^. Linkage disequilibrium analysis among SNPs was performed within a 100 kb window, 100 variant steps, and an R^2^ threshold of 0.2, resulting in 261,217 SNPs selected for PRS construction. These SNPs’ GWAS effect sizes were aggregated to compute the PRS using PRSice-2 for each individual and pain phenotype, incorporating only SNP effect sizes from summary GWAS data of the training sets.^30^ The PRS included all GWAS covariates and applied 20 P-value thresholds for SNP inclusion, ranging from P = 5×10^-8^ to P=1. Additionally, pathway-based PRS were analyzed using PRSet with Neural/Immune Gene Ontology (NIGO) pathways, focusing on those containing 10 to 100 genes for enhanced specificity.^31,32,33^

#### Brain imaging

A sub-sample of baseline participants were invited to attend a follow-up roughly 9 years later (range: 3-13 years), hereafter referred to as the 9-year follow-up. At the time of this study, data were available for 49,001 participants who had completed this follow up, which included a Magnetic Resonance Imaging (MRI) scan of the brain and the same questionnaires and assessments as baseline^3^. Structural and functional brain features were derived from three neuroimaging modalities, including T1-weighted MRI, diffusion MRI (dMRI) and resting-state functional MRI (fMRI). The image processing pipeline, artifact removal, cross-modality and cross-individual image alignment, quality control and phenotype calculation are described in detail in the central UK Biobank brain imaging documentation (https://biobank.ctsu.ox.ac.uk/showcase/showcase/docs/brain_mri.pdf) and by Alfaro-Almagro and colleagues^4^. Additionally, we excluded participants with excessive head motion or suboptimal signal-to-noise ratio, as well as those whose T1 brain images required excessive warping for non-linear alignment to standard space, defined quantitatively as deviations greater than three standard deviations from the group mean (exceeding the 99.75th percentile).

#### T1-weighted MRI

(1041 features, N = 41,979) T1-weighted MRI features include regional and subcortical gray matter volume, cortical thickness, surface area, and volume, regional gray/white matter intensity contrast as derived from T1-weighted MRI. Participants missing data for more than 75% of the features (exceeding 780 features) were excluded from further analysis, the remaining missing data was imputed using the median.

#### Diffusion-weighted MRI

(614 features, N = 36,779) Diffusion-weighted MRI features include microstructural measures of white matter tracts including mean fractional anisotropy and mean diffusivity as well as derivatives of diffusion tensor imaging including orbital diffusivity, MO, L1, L2, ICVF, and ISOVF. Participants missing more than 75% of the features (exceeding 461 features) were excluded from further analysis. This resulted in the final sample of 36,779 individuals with complete diffusion features, as such no imputation was required.

#### Resting state functional MRI

(38,781 features, N = 37,414) Resting-state functional Magnetic Resonance Imaging data is based on the minimally preprocessed pipeline designed and carried out by the FMRIB group, Oxford University, United Kingdom. The minimally preprocessed resting-state fMRI data from the UK Biobank were analyzed using the following preprocessing steps: motion correction with MCFLIRT^4^, grand-mean intensity normalization, high pass temporal filter, fieldmap unwarping, and gradient distortion correction. Noise terms were identified and removed using FSL ICA-FIX. Full information on the UK Biobank preprocessing is published^4^. Additional preprocessing included warping the image in native space to the 3mm MNI template (FSL), despiking using 3DDespike (AFNI from Nipype), 6-mm kernel smoothing (Nilearn), and resampling to 3-mm (for storage purposes). A modified Brainnetome atlas^5^ including additional midbrain, brainstem, and cerebellar regions was used to parcel the brain into 279 distinct regions. Dynamic connectivity was estimated between each region to derive a functional connectome using Dynamic Conditional Correlation (DCC). DCC is based on generalized autoregressive condition heteroscedastic (GARCH) and exponential weighted moving average (EWMA) models (implemented by https://cocoanlab.github.io/tops/).

#### Bone structure

(68 features, N = 40,815) At the time of this study, measures from Dual-energy X-ray Absorptiometry (DXA) bone scans were available for 40,815 participants at the 9-year follow-up (https://biobank.ndph.ox.ac.uk/showcase/ukb/docs/DXA_explan_doc.pdf). Radiographers analyzed the scans at the whole body, spine, and hip levels either during or shortly after the image acquisition. This analysis facilitated the generation of comprehensive numerical measures, including bone area, bone mineral content (BMC), and bone mineral density (BMD). A total 91 features across seven bone systems—spine, femur, head, legs, pelvis, arms, and ribs were extracted by the UK Biobank. From this pool, we excluded features with low coverage (fewer than 10,000 participants or 25% of the sample), narrowing the feature space to 68. Among the 40,815 participants assessed, we imputed any remaining missing data, which affected 4.9% or 2,015 values, using the median.

### Psychosocial Phenotypes (81 features, N = 493,211)

We used a truncated set of psychosocial features originally defined in our recent paper, see Tanguay-Sabourin, C. *et al* for details^6^. As the goal of the present study is to compare biological predictors to psychosocial predictors, we excluded biological features included in the original model to generate a strictly psychosocial modality, including BMI, age, sex, ethnicity, grip strength, and blood pressure. Thus, a total of 81 features collected at the baseline and follow-up visits were selected a-priori based on their relevance to chronic pain. The selection was based on the Prognosis Research Strategy (PROGRESS) group who recently provided a framework for the development of a prognostic model to determine risk profile^7^. Variables were organized through an iterative approach along a hierarchical framework from 81 features into 8 categories forming three distinct domains (i.e., mental health, physical health, and sociodemographic). The three domains are as follows:

#### Mental health

The mental health domain includes 3 categories: i) neuroticism – all individual items and their total sum-score – based on 12 neurotic behaviors closely linked to negative affect, ii) traumas – illness, injury, bereavement, or stress in the last 2 years – including 6 events, and iii) mood – reported frequency of certain moods in the past 2 weeks and visits to a general practitioner or psychiatrist for nerves, anxiety, tension, or depression.

#### Physical health

The physical health domain includes 3 categories: i) physical activity – Metabolic Equivalent Task (MET) scores computed using the International Physical Activity Questionnaire (IPAQ) guidelines^8^, ii) sleep – questions regarding duration, napping, snoring, and sleeplessness, iii) substance use – smoking and alcohol use.

#### Sociodemographic

The sociodemographic domain includes 2 categories: i) socioeconomic status – education completed, income, employment, etc., ii) occupational measures – individuals present within household, social entourage, and job type (e.g., manual or physical job).

For a detailed view of the biological and psychosocial features included in this study, see **Supplementary Tables 1-6.**

### Pain phenotypes in the UK Biobank

#### One-month pain

Participants reported if they experienced pain impacting their usual activities at any major anatomical body sites (head, face, neck or shoulder, back, stomach or abdominal, hip, knee, or pain all over (PAO)) in the last month. Participants answering PAO were not allowed to report additional pain locations. This category consists of both chronic and acute pain.

#### Acute and chronic pain sites

Participants reporting pain at a given site in the last month were then asked if the pain at that site had persisted for more than 3 months. This question was used to distinguish between a chronic pain site, one present for more than 3 months, and an acute pain site, one present for 3 months or less, according to the classification from the International Association for the Study of Pain^1^.

#### Deriving 14 target pain phenotypes

From these data, we derived 14 pain phenotypes, including a general chronic pain phenotype representing pain at any of the body sites, and a general acute pain phenotype. Additionally, we identified seven chronic pain site phenotypes, each representing pain experienced at one of the specified body sites, and four phenotypes that quantified the number of chronic pain sites reported, categorizing the extent of chronic pain spread ranging from 1 to 4 or more distinct sites.

### Pain-Associated Diagnoses

Participants’ diagnoses were sourced from both self-reported interviews conducted at UK Biobank assessment centers (Field IDs 20001 and 20002) and healthcare records provided by the UK National Health Services (NHS). In the interview process, trained nurses validated and, if necessary, refined the medical conditions that participants initially reported through a touchscreen questionnaire. In cases of uncertainty, participants described their condition to a nurse, who then assigned a suitable code or logged it as a free-text description. This free text was later matched to a specific entry by a physician.

For health outcomes resulting in hospital admissions, hospital inpatient records utilized the International Classification of Diseases and Related Health Problems (ICD-10) coded primary or secondary diagnoses (Field IDs 41270 and 41271). Meanwhile, primary care data, available for 45% of the study cohort (n = ∼230,000), were obtained from Read Codes v2, as coded by general practitioners (Field ID 42040). The UK Biobank additionally provided curated data fields indicating the first occurrence of a set of diagnostic codes (Category: 1712) for a wide range of health outcomes across self-report, primary care, hospital inpatient data and death data, mapped to a 3-digit ICD code. For broader diagnoses that were effectively captured with a three-digit ICD code (e.g., Rheumatoid arthritis; ICD code: M05 & M06), we extracted information from this first occurrences database. In contrast, for conditions defined by more granular ICD coding (e.g., ankylosing spondylitis; ICD code: M081), data were manually curated from self-report and hospital inpatient records.

For our analysis, we selected 35 pain-associated diagnoses based on their significant pain prevalence and ample sample size. Criteria included having over 45% pain prevalence (acute or chronic) and an occurrence exceeding 100 participants at the 9-year follow-up. These diagnoses were then aligned with the first occurrences database, ICD-10 codes and primary care data, collating all participants with a record for each specific diagnosis. For instance, the sciatica group amalgamated both self-reported and healthcare recorded (ICD codes: M543 & M544) instances of sciatica. A detailed list of codes for the 35 diagnoses is available in **Supplementary Table 7**. To ascertain that an illness’s onset or diagnosis predated a given study visit, we contrasted the earliest recorded illness date with the participant’s respective visit date at each visit (e.g. baseline, 4-year, and 9-year follow-up).

To enable comparisons across diagnoses, we defined the healthy control group as participants with no self-reported diagnoses and no self-reported or healthcare recorded instances of the 35 pain-associated diagnoses. This resulted in a healthy control sample size of 103,034 (20.5% of full sample size) participants at the baseline visit and 5,237 (11.6% of full sample size) participants at the 9-year follow-up. As such, the control population may not be representative of the full study population.

Demographic information including sex, ethnicity, and age distributions for each diagnosis within the UK Biobank, is detailed in **Extended Data Fig. 4**.

### Validation cohorts

To validate the biomarkers identified in this study, data were sourced from two distinct validation cohorts: the All of Us Research Program (AoU) and four datasets from the OpenPain repository (**Extended Data Fig. 8**).

#### All of Us Research Program

All of Us Research Program aims to enroll over a million U.S. participants, with a focus on individuals from underrepresented groups in research. The program collaborates with healthcare organizations to collect and share electronic health records (EHR) from consenting participants, which includes any healthcare diagnosed medical conditions and blood assays the participants have had collected. These records are standardized to the Observational Medical Outcomes Partnership (OMOP) Common Data Model by data specialists at each organization, enabling uniformity across different EHR systems^9^.

#### OpenPain repository datasets

OpenPain is a repository featuring data from brain imaging studies on chronic pain. We utilized four studies from OpenPain that provided whole-brain resting-state fMRI imaging for chronic pain subjects and pain-free controls. The first dataset involved UK participants with chronic back pain (24 patients, 30 controls), the second dataset comprised Japanese chronic back pain patients (17 patients, 17 controls), the third and fourth datasets featured US patients with chronic back pain (74 and 66 patients, respectively, with 22 controls in the fourth dataset). These datasets were amalgamated to form a unified validation cohort totaling 181 patients and 69 controls. Consistent brain imaging processing protocols were applied across all datasets, as described below:

Preprocessing was conducted using fmriprep version 1.4.1, incorporating the following preprocessing steps: motion correction with MCFLIRT, susceptibility distortion correction to correct for field inhomogeneities, registration to the T1w image using FLIRT, followed by warping from native space to the 3mm MNI template (FSL), and removal of physiological and motion-related noise terms, including raw signals, squared signals, and their first derivatives for white matter, cerebrospinal fluid, translations, and rotations along x, y, and z planes from timeseries using Nilearn’s signal.clean function. Aligning with the preprocessing of the UK Biobank fMRI data, we included connectivity despiking using 3DDespike (AFNI), 6-mm kernel smoothing (Nilearn), and resampling to 3-mm, followed by parcellating the brain into 279 distinct regions using the Brainnetome atlas and estimating parcel-wise dynamic functional connectivity using Dynamic Conditional Correlation (DCC). Lastly, neurocombat harmonization was employed to mitigate site-specific effects in the data.

## Statistical analysis

### Machine learning Models

Predictive machine learning models were constructed to classify the 14 pain phenotypes and 35 pain-associated diagnoses from pain-free or diagnosis-free controls, respectively. Machine learning models implemented nested cross-validated logistic regression (implemented using SnapML) with a randomized hyperparameter search (implemented using scikit-learn) to optimize the ridge regression regularization hyperparameter (‘l2’) for each individual model. To address cases of class imbalance, the ’class_weight’ parameter in the models was set to ’balanced’. This adjustment modifies the loss function penalty to ensure that the majority class does not disproportionately influence the model. Both inner and outer layers of the nested Cross-Validation (CV) utilized a 5-fold strategy (see **Extended Data Fig. 1** for a schematic of the modeling pipeline). To prevent data leakage and minimize model bias, feature preprocessing steps including standardization and residualization were fit to the training folds and then applied to the validation fold within each nested CV. Alternative algorithms such as support vector machines and gradient boosting trees were evaluated but were not chosen as the primary machine learning methodology based on their performance metrics (**Extended Data Fig. 2**).

Machine learning models were trained separately on features comprising the 5 distinct modalities (blood, genetics, brain, bone, and psychosocial) to classify the 14 pain phenotypes and 35 pain-diagnoses. This approach yielded 70 distinct pain phenotype models (14 pain phenotypes multiplied by 5 modalities) and 175 distinct pain-diagnosis models (35 diagnoses multiplied by 5 modalities). Additional models were trained on the modality subcategories (inflammatory/immune, metabolic, hematological, T1 imaging, diffusion imaging, and resting state functional connectivity), for each pain phenotype and diagnosis.

To bolster the reliability of our findings and account for potential variability, each model underwent 5 iterations with unique random states (e.g., 5-times repeated 5-fold nested cross-validation). This procedure randomized the distribution of subjects across the training and test sets, from which we calculated confidence intervals for model performance metrics.

#### Mitigating confounding variables

The influence of confounding variables on machine learning predictions, especially in the context of biological data, is well-documented^11,12,13^. To mitigate potential biases introduced by confounders, we integrated regression-based deconfounding (i.e., residualization) within each of our modeling pipelines.

For each cross-validation fold:

1. A linear regression model was fit within the training data on features from a given modality to predict confounding variables.
2. This trained model was then applied to the features of both the training data and the left-out validation data to extract residual values, with the linear effects of the confounding variables removed. The residual values were then used as the feature space for model training (on training folds) and evaluation (on validation fold) of pain endpoints.

For a comprehensive list of the confounders addressed and their specific handling within each modality, refer to the detailed modality pipeline descriptions below:

#### Blood biochemistry pipeline

A significant portion of participants within the diagnosis groups reported taking medications known to alter blood biochemistry. These include, but are not limited to, hypertensive drugs, anti-rheumatic medications, and immunosuppressants. Recognizing the potential biases these medications could introduce in the biochemical features under study, we incorporated propensity score matching to control for them. This approach aimed to align subjects from the disease groups with their counterparts in the control group based on self-reported medication intake.

While unable to eliminate all confounding effects of medications, our method effectively mitigated a considerable extent of the confounding impact, as evident from **Extended Data Fig. 6**. To provide a brief overview:

- **Propensity Score Estimation**: Propensity scores, which represent the likelihood of an individual being treated considering specific confounds, were generated via a logistic regression model. We extracted confounds by implementing the top 5 components from a mixed factor analysis on the predominant 5 medications, including statins, associated with each disease group. This, combined with age and sex, served as the foundation for our matching process between case and control groups.
- **Matching Process**: Each individual from the case group was paired with a counterpart in the control group using the Hungarian matching algorithm^14^. This algorithm determines an optimal match by minimizing the overall distance or "cost" between paired subjects. This distance is calculated based on the absolute difference in their estimated propensity scores. A conservative caliper, defined as 0.2^11^, ensures that this difference does not surpass a predetermined threshold.
- **Timing and Application**: It’s important to note that propensity score matching was executed prior to the cross-validation data split. This ensured consistency in the data across every modeling iteration.

#### Polygenic risk score pipeline

In the polygenic risk score models, we applied a thresholding procedure to the outputs of each diagnosis GWAS, yielding 20 levels of statistical significance thresholds for each single nucleotide polymorphism, ranging from P = 5×10^-8^ to P=1. Models were subsequently trained on these thresholds to determine the optimal P-value threshold to distinguish between the pain outcome group and the healthy control group, without adjusting for any confounding variables.

#### Brain imaging pipeline

Brain imaging models were trained on multi-modal brain imaging features including white matter features from diffusion-weighted imaging, structural features from T1-weighted imaging, and functional connectivity features from resting state imaging. Several confounding variables were adjusted for in brain imaging pipelines: average absolute head motion, volumetric scaling (to account for variations in brain size) from the T1 image to a standardized space, age, and sex.

#### Bone structure pipeline

Bone structure models were trained on DXA derived features including bone area, bone mineral content (BMC), and bone mineral density (BMD), and were adjusted using age and sex as confounding variables.

#### Psychosocial pipeline

To align with the methodologies employed in other modalities we adjusted for age and sex as confounding variables in our models that were trained on psychosocial features.

### Classification of pain phenotypes

The area under the receiver operating characteristic curve (ROC-AUC) scores were calculated from models trained to differentiate between pain phenotypes and pain-free controls (Fig. 4 B, C, and D). ROC-AUC scores were obtained from the testing folds of five iterations of machine learning models, each employing a 5-fold cross-validation (CV) strategy, thereby yielding a total of 25 testing metrics per pain endpoint. The reported ROC-AUC scores estimate the effectiveness of the models in distinguishing between participants with a given pain phenotype (i.e. general chronic pain, general acute pain, chronic pain site, or number of chronic pain sites) and those without. Confidence intervals at the 95% level were calculated using 1,000 bootstrap resamples of the 25 AUC metrics. Additionally, heritability estimates were derived from GWAS of the general chronic pain, acute pain, and chronic pain site models.

### Phenotyping and classification of pain-associated diagnoses

Pain-associated diagnoses were phenotyped based on their self-reported chronic pain prevalence and segmented according to the number of reported pain sites, ranging from 1 to 4 or more. Furthermore, the localization of chronic pain for each diagnosis was determined by calculating the prevalence of pain at the 7 recorded body sites for each condition. These prevalence rates were then normalized (z-scored) across conditions for each specified site, providing a standardized measure of pain localization or distribution for each diagnosis (Fig. 1B).

The classification performance for classifying pain-associated diagnoses from diagnosis-free participants was assessed using the same methodology applied for the pain phenotype models. This approach resulted in 25 testing metrics for each diagnosis across each modality-specific machine learning model. To identify the most effective biological modality for classifying each diagnosis, we selected the modality with the highest average AUC score across modeling iterations for each diagnosis. We then computed 95% confidence intervals for these selected scores using 1,000 bootstrap resamples, with the results depicted in Fig. 1C. Adjacently, AUC scores derived from the psychosocial classification models are presented, enabling a side-by-side evaluation of the biological and psychosocial modalities’ performance across diagnoses. Comprehensive AUC scores for diagnoses across all modalities are provided in **Extended Data Fig. 5**. To interpret the distinct contributions of each modality to diagnosis classification, we trained additional models on modality subcategories. Consequently, AUC scores were derived from models trained on the blood modality subcategories, including inflammatory/immune, metabolic, and hematological markers. For the brain modality, models were trained on T1 structural features, diffusion-weighted imaging for white matter integrity, and resting-state functional connectivity. Within the psychosocial modality, we investigated eight subcategories: mood, neuroticism, life stressors, sleep quality, substance abuse, physical activity levels, socioeconomic status, and occupational factors. However, the bone structure and genetic modalities remained undivided and were analyzed as whole. Within diagnosis averaged AUC scores obtained from the subcategories—or from entire modalities in the cases of bone structure and genetics—were subsequently standardized using z-scoring relative to other diagnoses within the same modality or subcategory. By z-scoring the AUC values, we aimed to delineate whether certain diagnoses are associated with unique biological and/or psychosocial alterations or if they reflect broader deviations across multiple domains of biological and/or psychosocial functioning. This approach allowed a comparison of how pain-associated diagnoses are characterized by distinct or overlapping biological and psychosocial profiles.

The deviance explained by biological factors, psychosocial factors, and their unions in classifying two distinct chronic pain conditions - rheumatoid arthritis and fibromyalgia - was assessed using the optimal biological modalities for each condition, determined by the highest average AUC scores: blood for rheumatoid arthritis and brain for fibromyalgia. These calculations were then compared with the deviance explained in models of self-reported chronic pain, employing the same biological modalities for comparability.

The deviance explained (D^2^) by the biological (B), psychosocial (P), and combined (B+P) predictors was calculated following a modified approach based on the methodology outlined by Dinga et al. 2020, adapted to our context of comparing different types of predictive information rather than controlling for confounds^12^. First, predicted probabilities were extracted from models trained on: biological predictions (B) from the blood or brain models, psychosocial predictions from the models trained on psychosocial data (P), and combined data (B+P) integrating both biological and psychosocial predictions.

Logistic regression models were then fit on these predicted probabilities to predict each specified outcome (either the diagnosis or self-reported chronic pain), generating:

- A model using only predicted probabilities from biological data (B).
- A model using only predicted probabilities from psychosocial data (P).
- A model using combined predicted probabilities from both data sources (B+P).

The deviance for each predictive model was then calculated using the following formula:

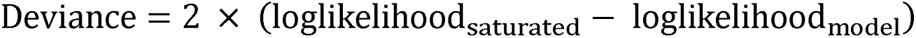

Where the saturated model represents a model with perfect fit to the data and the maximum possible log-likelihood. Subsequently, the fraction of deviance explained (D^2^) for each model was derived by normalizing the model’s deviance against that of a null model, which includes only an intercept, according to:

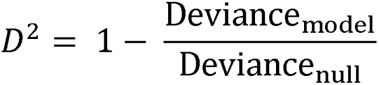

To dissect the deviance explained into unique and shared contributions, the following calculations were performed:

- 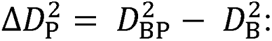 The unique contribution of psychosocial predictions beyond what is explained by biological predictions.
- 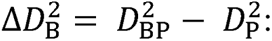 The unique contribution of biological predictions beyond what is explained by psychosocial predictions.
- 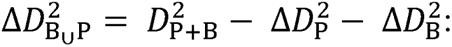 The shared or overlapping contribution of biological and psychosocial predictions to the explained deviance.

Where 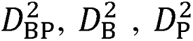 are deviance explained of models containing biological and psychosocial predictions, biological predictions, and psychosocial predictions, respectively. These results quantify the extent to which biological and psychosocial factors individually and jointly explain the variance in the binary disease or self-reported pain outcome and are displayed in **Fig. 1D**.

### Nociplastic Functional Signature (NFS)

We developed a predictive resting-state functional signature for nociplastic pain conditions within the UK Biobank cohort at the 9-year follow-up. Based on established literature, we identified diagnoses categorically defined as nociplastic conditions, which included fibromyalgia, chronic fatigue syndrome, and widespread chronic pain^15,16,17^. Nociplastic conditions are believed to be mechanistically linked to central sensitization or alterations in central nervous system processing of pain leading to symptoms such as diffuse pain, fatigue, sleep disturbances, and cognitive impairments^16,18,19^.

To develop the Nociplastic Functional Signature (NFS), we combined fibromyalgia, chronic widespread pain, and chronic fatigue syndrome into a single nociplastic condition (n = 535) and entered them into our machine learning pipeline to differentiate from diagnosis-free participants based on resting-state functional connectivity data. Structure coefficients were then derived from the predictive model to create the signature. These coefficients highlight the connectivity patterns most predictive of the nociplastic condition, and are described below:

#### Structure coefficients

Given the potential instability and bias in feature weights derived from multivariate predictive models, we utilized structure coefficients, or model encoding maps, to identify features individually linked to the outputs of our predictive machine learning models^20^. Structure coefficients, recognized in brain imaging and predictive modeling research^21,22^, provide a means to extract reliable features associated with a predicted outcome. Specifically, these coefficients reveal the individual association of features—in this case the Dynamic Conditional Connectivity (DCC) between two brain regions of interest (ROIs)—with the model’s output—in this case the predicted probability of a nociplastic pain condition—thereby mapping individual features to the overall multivariate model prediction.

For each participant, structure coefficient maps were generated by computing the covariance between the predicted probabilities from the logistic regression model and the parcel-wise DCC values. This process created an encoding map delineating the directionality of the relationship between each feature and the model prediction. In our analysis, this method maps which brain voxels were positively or negatively associated with the predicted nociplastic condition. For all predictive models, structure coefficient maps were calculated for each of the five iterations of the machine learning pipeline and subsequently averaged.

#### Measuring functional dysconnectivity in nociplastic conditions

Structure coefficient maps, derived from models individually trained to predict fibromyalgia, widespread chronic pain, chronic fatigue syndrome, were correlated using two-sided Pearson’s coefficients, with significance levels adjusted for multiple comparisons using the Bonferroni method. To highlight key cortical regions involved in each condition, these maps were node-averaged and thresholded to identify the top 25% of absolute coefficient values and visualized on cortical surface renderings. It is important to note that the correlation analyses between condition coefficients utilized the complete, unthresholded coefficient vectors, which included both subcortical and brainstem areas. However, the visual representations focused solely on the top 25% of coefficients within cortical regions of interest (ROIs) as shown in Fig. 3B. The same approach was taken to visualize the NFS in Fig. 3C. Surface rendering was performed using the Surf Ice software, available at https://www.nitrc.org/projects/surfice/.

#### Validation of the NFS

The NFS was validated in four external datasets available in the OpenPain repository. These datasets were merged to form a validation cohort of 250 participants, allowing for an evaluation of the NFS’s generalizability in classifying chronic pain. This performance was then benchmarked against the Tonic Pain Signature (ToPS), a previously validated marker for sustained experimental pain^23^. ToPS weights were applied to the participant-level DCC data (brainnetome atlas parcellation) in the OpenPain dataset across various thresholds (100%, 50%, 25%, 10%, 5%, 1%, 0.5%) to test for generalizability. The same thresholding procedure was employed with the NFS to establish its performance within the OpenPain dataset. For each threshold, we calculated the AUC to differentiate between participants with chronic pain and pain-free individuals. A null AUC distribution for each threshold was generated by randomizing the outcome variable across 1,000 permutations and recalculating the AUC (Fig. 3E).

### Composite blood assay signature

A composite blood assay signature capturing the common alterations in blood assay features across well-predicted diseases was created. Included were 13 diseases for which blood assay models predicted at a ‘good’ or better performance level (AUC >= .70). This signature was formed by calculating the average of the structure coefficients from the predictive models for each of the 13 diseases. The resulting map represents the direction and magnitude of the 52 blood assay features that are consistently altered across these conditions (Fig. 2C). The signature could then be transformed into a subject-level risk score by taking the dot product of the signature and a subjects standardized and de-confounded blood assay feature profile.

#### Assessment of composite blood assay signature

The blood assay signature was assessed by testing its ability to predict cross-sectional disease prevalence at baseline and longitudinal disease incidence at roughly 4 and 9 years in the UKB. First, subject-level signatures were derived by taking the dot product of the signature coefficients and each subjects standardized and de-confounded (residualization using age and sex as confounds) blood assay feature profile at baseline. For each of the 13 diseases included in the risk score, subjects were divided into 3 case groups: 1) disease present at baseline 2) disease onset at 4 year visit 3) disease onset at 9 year visit. Similarly, 3 control groups were created 1) diagnosis-free at baseline 2) diagnosis free at 4 year visit 3) diagnosis free at 9 year visit. Each case group was matched with its respective control pairing and effect size estimates were calculated between the risk scores for the pairs using Cohens d (95% confidence interval derived from 1,000 bootstrap iterations) and AUC scores for discrimination performance. Importantly, the control group within each timepoint analysis remained the same across diseases (Fig. 2B).

The signature was also evaluated on a pooled diagnoses phenotype, where subjects with any of the 13 diagnoses were aggregated into a single disease group at each timepoint. The pooled diagnosis group was then compared to its timepoint-matched control group to calculate effect sizes, using the same methodology as described previously for the individual diagnosis (Fig. 2D).

#### Disease progression analysis

We analyzed repeated blood assay data from the 4-year follow-up (n = 19,360) to measure signature evolution over time. The composite signature coefficients were applied to the repeat blood assay data, yielding a follow-up signature. This analysis included the three diagnosis case groups (e.g., disease present at baseline, disease onset at 4 year, disease onset at 9 year) to examine signature dynamics across varying disease progression stages relative to control subjects with no diagnoses at follow-up. We employed a linear mixed effects model to analyze changes in the signature from baseline to follow-up, incorporating fixed effects for time, disease group, and their interaction, and accounted for individual variability by considering participants as a random effect. We used Bonferroni correction to assess the significance of group-time interaction terms for signature changes between each disease group and healthy controls (Fig. 2E).

### Validation of composite blood assay signature

#### Validation in the All of Us Research Program (AoU)

In the All of Us Research Program (AoU), a simplified version of the composite blood signature was developed to fit the program’s available assays, resulting in a simplified signature with 10 retained assays detailed in **Supplementary Table 1**. This validation involved defining pain-diagnosis and control groups through curated ICD-10-CM codes within the AoU’s Dataset and Cohort Builders, selecting individuals based on their recorded diagnoses of the 13 conditions relevant to the original signature. Healthy controls were defined as participants without inflammatory, musculoskeletal, or cardiovascular diagnoses. We applied this sparse signature to AoU participant blood assay profiles, adjusting for age and sex, to create participant-level sparse signatures. The effectiveness of this simplified signature was quantified using Cohen’s d effect sizes and AUC scores, derived from comparisons across individual diagnosis groups and controls, incorporating 95% confidence intervals from 1,000 bootstrap iterations (Fig. 2E & F).

### Biological and psychosocial risk scores

For the blood, brain, and bone modalities, we identified diagnoses with sufficient classification accuracy, defined by AUC values greater than approximately 0.70. Only one disease was predicted 0.70 AUC or greater using the polygenic risk score modality, so it was excluded from this analysis. This approach yielded 13 diagnoses for the blood modality, 5 for the brain, and 4 for the bone. Within each modality, we generated biological and psychosocial risk scores for both diagnosed participants and diagnosis-free controls. These scores were calculated by averaging the log-transformed predicted probabilities from the respective disease prediction models. For instance, in the brain modality, for participants diagnosed with any of multiple sclerosis, stroke, fibromyalgia, chronic fatigue syndrome, or cervical spondylosis, we averaged the predicted probabilities from each brain model to establish a common biological risk score. Similarly, we computed a common psychosocial risk score by averaging the predicted probabilities from the psychosocial models for these conditions. In each modality group, participants were sorted into quintiles based on their biological and psychosocial risk scores, categorizing them into five risk levels: Low, Reduced, Neutral, Elevated, or High (Fig. 5A).

#### Biopsychosocial synergy in disease

In the blood modality analysis, we computed odds ratios for each of the 13 diagnoses independently against diagnosis-free controls. We determined the odds ratio using the unconditional maximum likelihood estimate, comparing the odds for participants within a specific quintile (e.g., ‘Low’) to those of all other participants in the cohort. This approach aimed to quantify the likelihood of having a specific condition (e.g., gout) based on a participant’s placement in each of the biological and psychosocial risk quintiles. Next, we calculated odds ratios for each diagnosis based on combinations of biological and psychosocial quintiles, assessing the likelihood of a diagnosis for participants categorized within both ’Low’ biological and psychosocial quintiles (LL), ’Reduced’ (RR), and so forth, up to the ’High’ (HH) combination (Fig. 5A).

Logarithmic odds ratios were computed for pooled (combined) diagnoses within each modality, covering all 25 possible combinations of biological and psychosocial risk quintiles. For each modality, Cohen’s d effect sizes were also calculated, comparing common risk scores for each diagnosis against scores from diagnosis-free individuals (Fig. 5B, C, D).

#### Longitudinal analysis of biopsychosocial synergy

Within the blood modality, we analyzed the onset of pooled diagnoses at the 4-year follow-up based on baseline biopsychosocial risk quintiles. We calculated log-odds ratios for each of the 25 combinations of baseline biological and psychosocial risk quintiles, comparing those with new onset diagnoses (n = 1,006) at follow-up to those who remained diagnosis-free (n = 1,282), as presented in Fig. 5F.

The incidence of a diagnosis within a 15-year period following baseline assessment was analyzed across four groups categorized by their baseline biological and psychosocial risk quintiles: Low–Low, High–Low, Low–High, and High–High. Kaplan–Meier curves were employed to visualize the time to diagnosis across these groups. Additionally, Cox proportional hazards models provided hazard ratios and p-values, while log-rank tests were applied to identify significant differences in the hazard rates, particularly comparing the High–High and Low–High groups (Fig. 5G). Survival analyses we’re conducted using the lifelines package in Python.

### Biopsychosocial chronic pain pathway

A structural equation model (SEM) was employed to investigate the relationships between biological and psychosocial markers and their effects on the development of rheumatoid arthritis and subsequent pain outcomes. Logarithmic predicted probabilities derived from the blood and psychosocial models of rheumatoid arthritis served as baseline biological and psychosocial risk indicators. An interaction term for the biopsychosocial interface was derived by multiplying these biological and psychosocial risks.

Pain outcomes were measured using the UK Biobank’s online pain questionnaire, which assessed pain across three dimensions:

1. Pain Impact: Assessed with the Brief Pain Inventory (BPI-39), the functional impact of pain was evaluated across seven areas: general activity, mood, walking ability, work, interpersonal relations, sleep, and life enjoyment, each on a scale from 0 (no interference) to 10 (complete interference). This measure was specifically targeted at participants reporting chronic pain at particular body sites.^24^
2. Pain Spread: The extent of pain was quantified by asking participants if they experienced pain or discomfort persistently or intermittently over more than 3 months, followed by specifying the body sites affected in the last three months. A summative phenotype representing the spread of pain was created based on the number of reported pain sites.
3. Pain Intensity: Chronic pain sufferers were prompted to rate their most bothersome pain at its worst in the past 24 hours on a scale from 0 (no pain) to 10 (pain as severe as imaginable).

The model specification was delineated as follows: rheumatoid arthritis development was modeled as a function of blood risk, psychosocial risk, and their interaction, while pain outcomes were modeled as a function of rheumatoid arthritis development, blood, and psychosocial risk. Model fitting involved estimating parameters that best reflected the covariances among the observed variables. The fit of the model was assessed using standard indices, including the Comparative Fit Index (CFI), Root Mean Square Error of Approximation (RMSEA), and the Standardized Root Mean Square Residual (SRMR), to evaluate how well the model represented the data structure (Fig. 6). SEM were constructed using semopy.

## Extended data

### Biological amplification in pain spreading

We derived biological signatures from models predicting the number of self-reported pain sites, ranging from 1 to 4 or more, utilizing structure coefficients from predictive models. In blood modality analyses, we derived structure coefficients for each model to reveal blood assay signature similarity as pain increases in spread. For brain modality studies, we node-averaged the structure coefficients from resting-state connectivity data and visualized the top 25% of absolute values on cortical surfaces. In the bone modality, we averaged coefficients related to bone density, mineral content, and area across different skeletal regions—including the spine, femur, head, legs, pelvis, arms, and ribs. In genetic analyses, we identified the top 5% FDR-corrected Neuro-Immune Gene Ontology (NIGO) pathways from each PRS model specific to the number of pain sites and categorized these pathways into biological processes using REVIGO (**Extended Data Fig. 3**)^25^.

### Cross-prediction models

Cross-prediction models were employed wherein models initially trained to classify specific diagnoses were tested on alternative diagnoses they were not originally trained to identify. We extracted ROC-AUC scores, sensitivity, and specificity from these cross-prediction tasks. Sensitivity and specificity scores were averaged for each original diagnosis across predicted alternative diagnoses within both biological and psychosocial modalities (**Extended Data Fig. 6**).

### Impact of medication use on the composite blood assay signature

To assess medication impact on the blood assay signature, we considered 11 medication families linked to the 13 diagnoses comprising the signature (**Extended Data Fig. 7**). A network model mapped chi-squared associations between diagnoses and medication families, with edge betweenness clustering identifying diagnosis-medication clusters (**Extended Data Fig. 7**).

We then recalculated effect sizes and discrimination metrics for the signature, excluding patients taking individual medication families. For instance, statistics were re-evaluated after excluding patients on antiepileptic drugs (ATC code: N03A). This exclusion was systematically applied to each medication class to assess the signature’s predictive accuracy devoid of medication influences (**Extended Data Fig. 7**).

## Statistical analysis

Data pre-processing and statistical analyses were performed using Python v.3.8 (including Numpy (v.1.22.0), Pandas (v.1.4.3), Scipy (v.1.10.1), Sklearn (v.1.3.2), Nilearn (v.0.10.0), Lifelines (v.0.26.4), Semopy (v.2.3.9), and SnapML (v.1.9.1)). Nested five-fold cross-validation was used to obtain unbiased model performance results. Permutation tests (with 1,000 iterations) were used to test whether the associations by Pearson’s r correlation were significantly higher than a null association. We used bootstrap resampling with 1,000 iterations to indicate the estimated error in the Cohen’s d and ROC-AUC effect sizes. In all analyses, significance was based on PL < 0.05 for single testing and BonferonniL< 0.05 for multiple testing. Further details of the statistical methods are specified in each relevant section above.

## Ethical approval

The UKB was approved by the National Information Governance Board for Health and Social Care and the National Health Service North West Multicenter Research Ethics Committee (ref. no. 06/MRE08/65). All participants gave written, informed consent and the study was approved by the Research Ethics Committee (no. 11/NW/0382). Further information on the consent procedure can be found at https://biobank.ctsu.ox.ac. uk/crystal/field.cgi?id=200. Informed consent for participants in the All of Us Research Program was obtained either in person or via an eConsent platform, encompassing primary consent along with HIPAA Authorization for the research use of electronic health records (EHRs) and additional external health data. For each dataset acquired from the OpenPain repository, participants provided written informed consent that authorized the collection of brain imaging, behavioral data, and questionnaire responses. Protocols, consent forms and study procedures were approved by McGill Institutional Review Board and/or Douglas Mental Health University Institute Research Ethics Board. This study received ethics approval under IRB application number: A03-M20-21B (21-03-079).

## Data availability

All data provided from the UKB are available to other investigators online upon permission granted by www.ukbiobank.ac.uk. Restrictions apply to the availability of these data, which were used under license for the current study (project ID 20802). The All of Us Research Program data is accessible for individuals at approved institutions. Details can be found at https://www.researchallofus.org/register/. OpenPain data can be accessed openly at https://www.openpain.org/.

## Code availability

Detailed and annotated code will be available at GitHub (https://github.com/EVPlab). The medication classification performed by Wu et al.^34^ can be found in supplementary data from the original article (https://www.nature.com/articles/s41467-019-09572-5). Code to extract the ToPS by Lee et al.^23^ can be found online (https://cocoanlab. github.io/tops/).

