## Extended Data for "A Biomarker-Centric Framework for the Prediction of Future Chronic Pain"

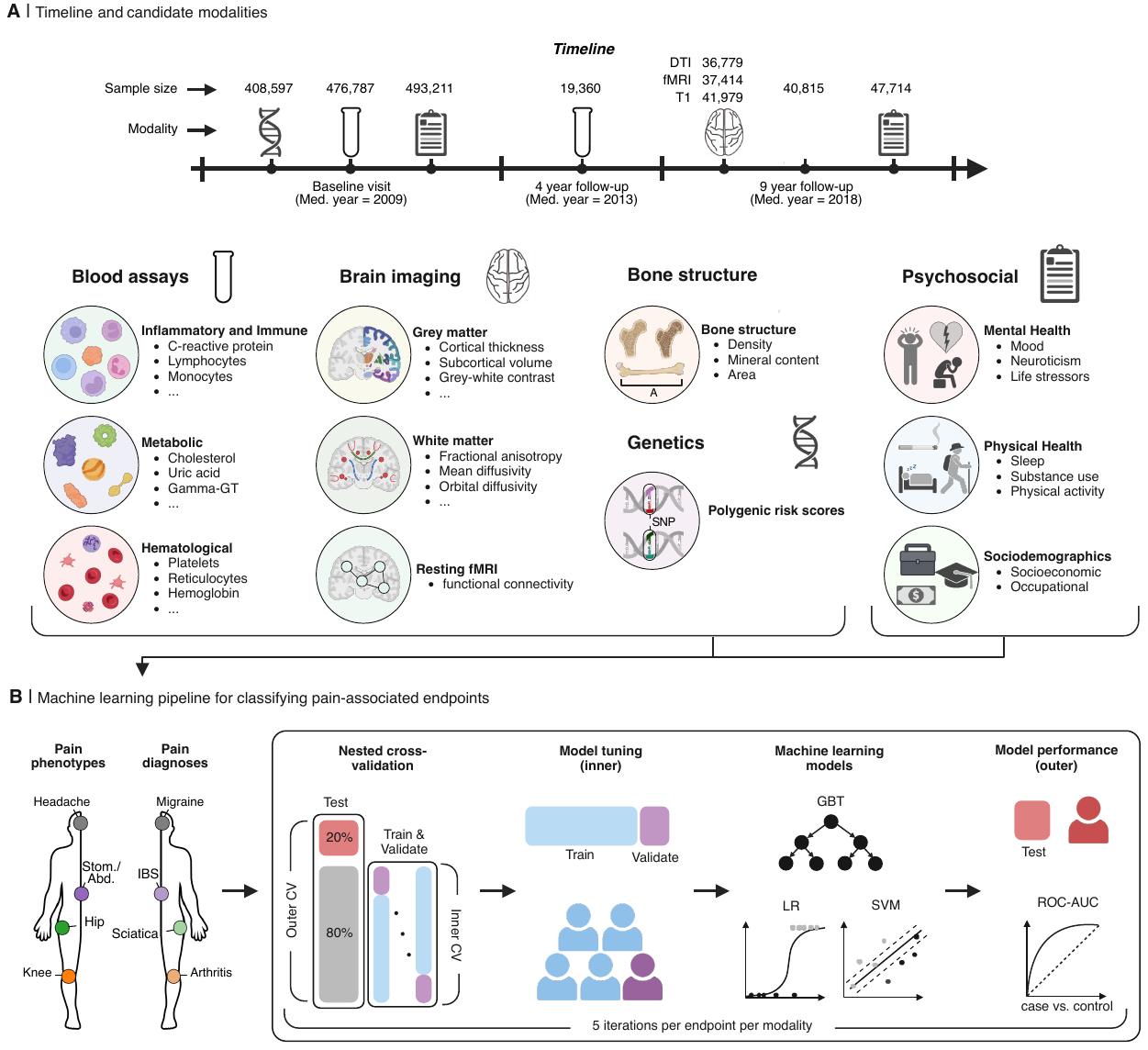

**Extended Data Figure 1: Candidate modalities and machine learning pipeline. A.** Top: A timeline depicts the collection of biological and psychosocial modalities across 3 different UK Biobank study sessions, with the number of samples available for analysis after data cleaning indicated above each modality icon. Below: Each modality is broken down into subcategories where applicable, with a description of example measures within these subcategories. Modalities are highlighted in large, bold font; subcategories in smaller bold font; and example measures are listed in regular font with bullets. **B.** A schematic outlines the machine learning pipeline employed to evaluate the ability of candidate modalities to classify pain endpoints. A nested Cross-Validation (CV) approach with 5-fold inner and 5-fold outer CV was utilized to optimize model performance without data leakage. The inner loop optimizes performance by training a model on each training fold and tuning hyperparameters on the validation fold to maximize the score. In the outer loop, the model's generalizability is gauged by averaging the scores across left-out test sets. Three machine learning algorithms we're assessed: gradient boosting trees, logistic regression, and linear support vector machines. This process was iterated five times for each modality, with participant order randomized in each iteration to prevent model performance bias based on train/test participant arrangement. GBT, Gradient boosting trees; LR, Logistic regression; SVM, Support vector machine; ROC-AUC, Receiver operating characteristic area under the curve.for age distribution, both for the full cohort and stratified by each diagnosis.

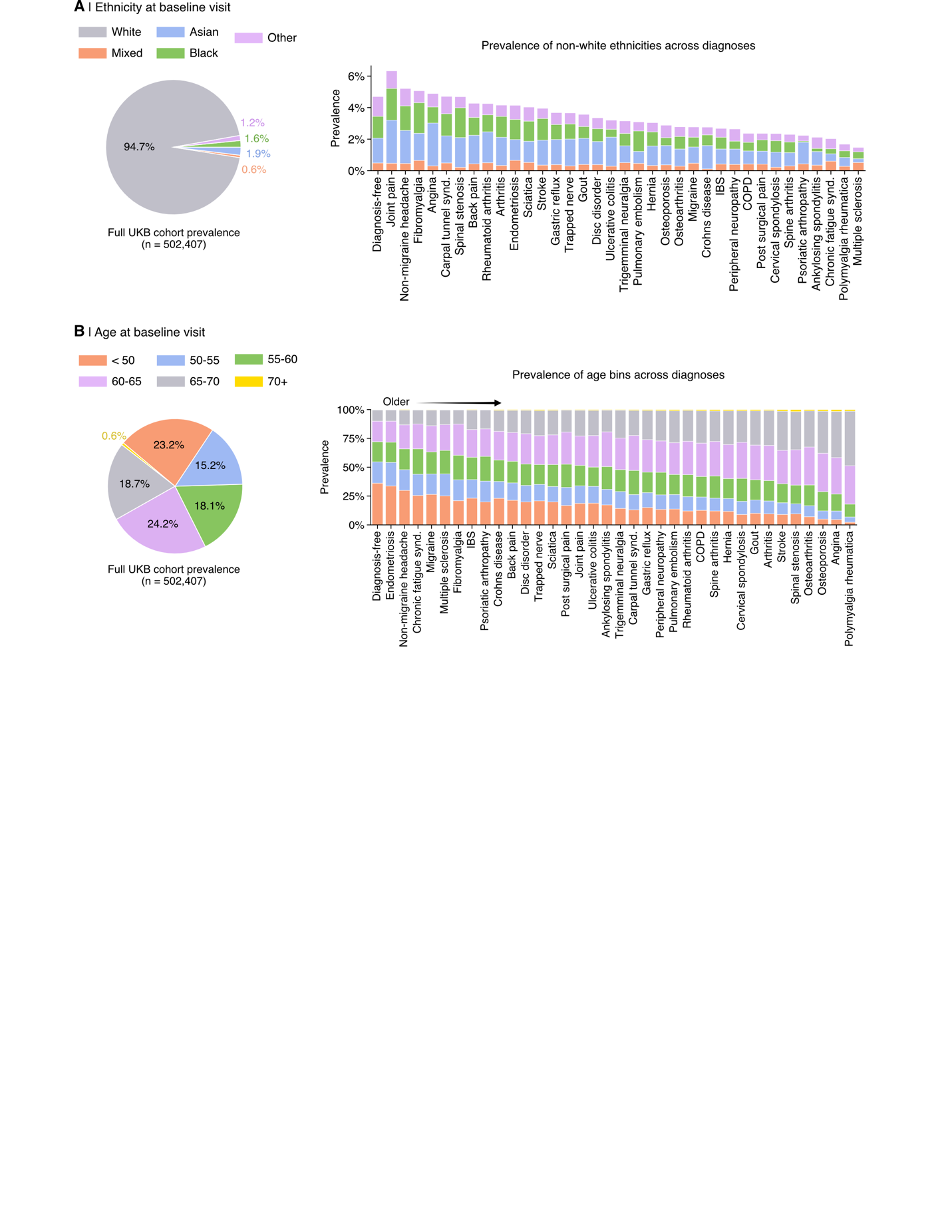

**Extended Data Figure 2: Ethnicity and age in the UK Biobank.** **A.** Ethnicity prevalence for the entire UK Biobank cohort (n = 502,407) is shown in the pie chart, with a breakdown of on non-white ethnicity prevalence across each pain-associated diagnosis shown using stacked barplots. In part **B**, data are visualized for age distribution, both for the full cohort and stratified by each diagnosis.

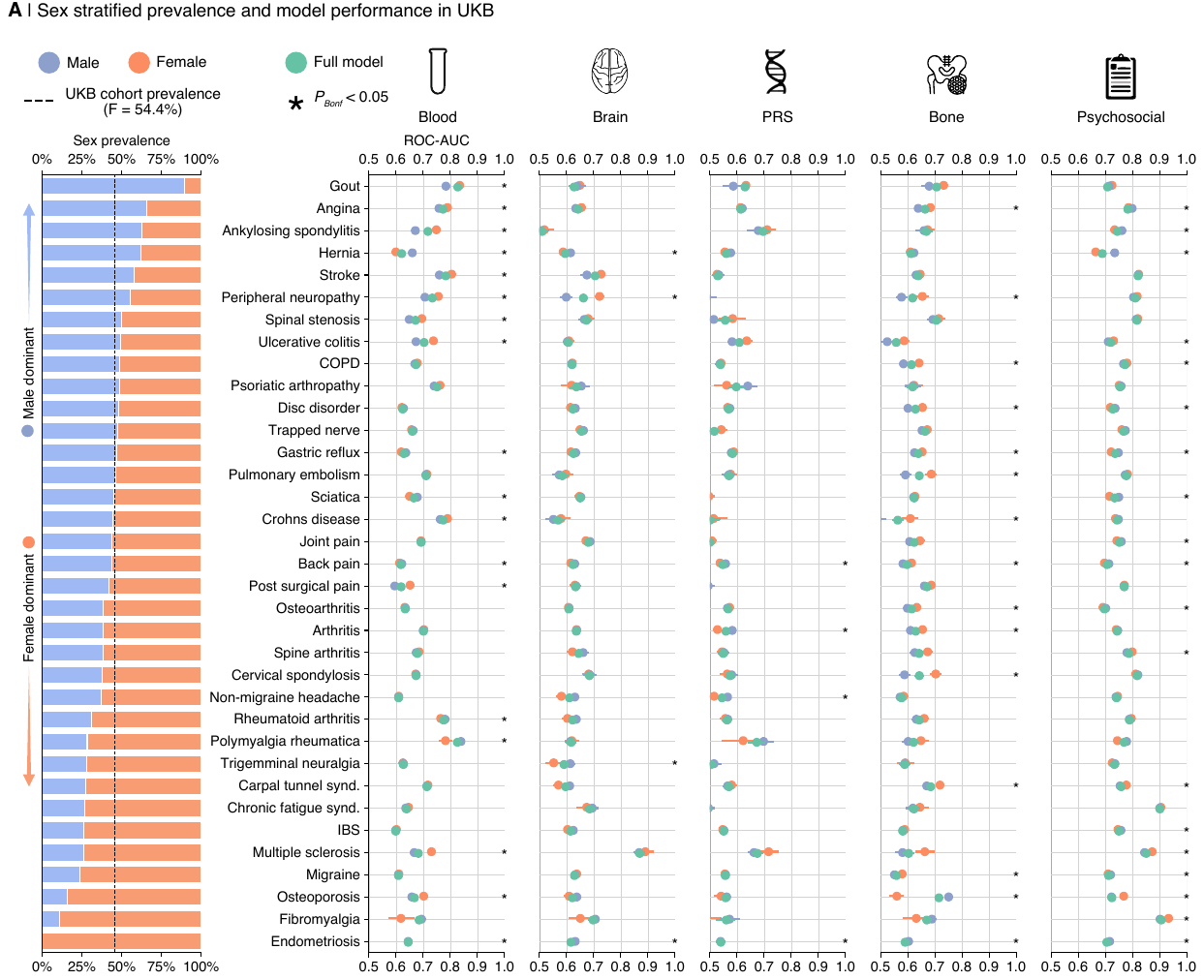

**Extended Data Figure 3: Sex-stratified performance in pain-diagnosis classification models.** The stacked barplot shows the sex prevalence for each pain-associated diagnosis assessed, with the overall UK Biobank cohort sex prevalence indicated by a dotted line. Models trained on both males and females were evaluated separately in males and females, using ROC-AUC scores from cross-validation testing folds. These scores depict the 95% confidence interval (1,000 bootstrap resamples) from 5 iterations of 5-fold CV for each model. Wilcoxon signed-rank tests assessed significant performance differences between sexes, with results Bonferroni corrected for multiple comparisons.

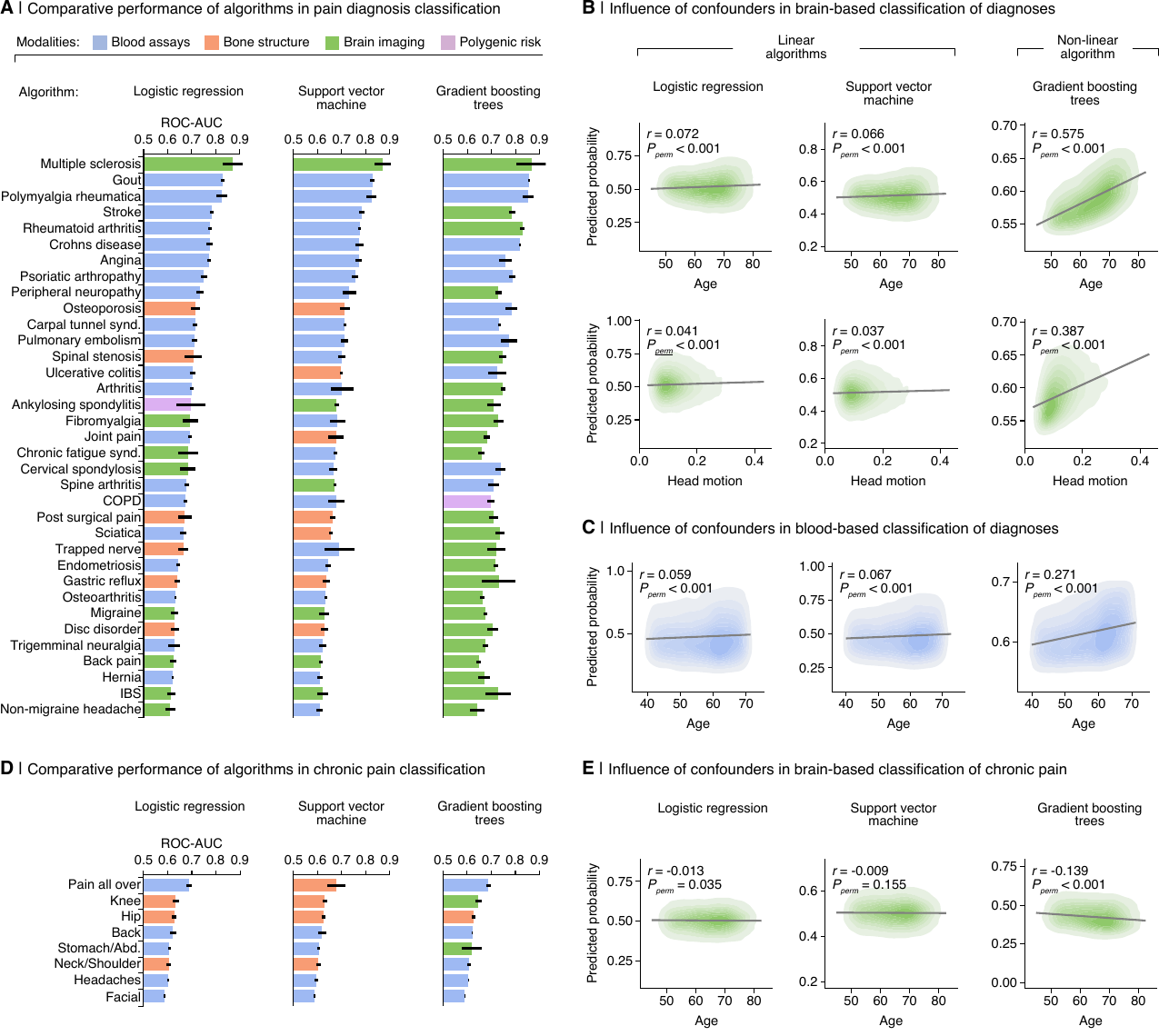

**Extended Data Figure 4: Evaluating alternative classification algorithms. A.** Receiver operating characteristic Area under the curve (ROC-AUC) scores depict the performance of three candidate algorithms in classifying pain-associated diagnoses. The ROC-AUC scores are colored corresponding to the modality that most accurately predicted each outcome, with results ordered according to the performance achieved by the logistic regression model. Regression density plots **B,C,E** show the association between the predicted probability of a diagnosis **B,C** or chronic pain site **E** (averaged across all models within a given modality) and confounds including age and/or head motion. Performance is also show for self-reported chronic pain body sites in **D**. These associations are quantified using two-sided Pearson's r correlations, with significance determined through 1,000 permutation tests.

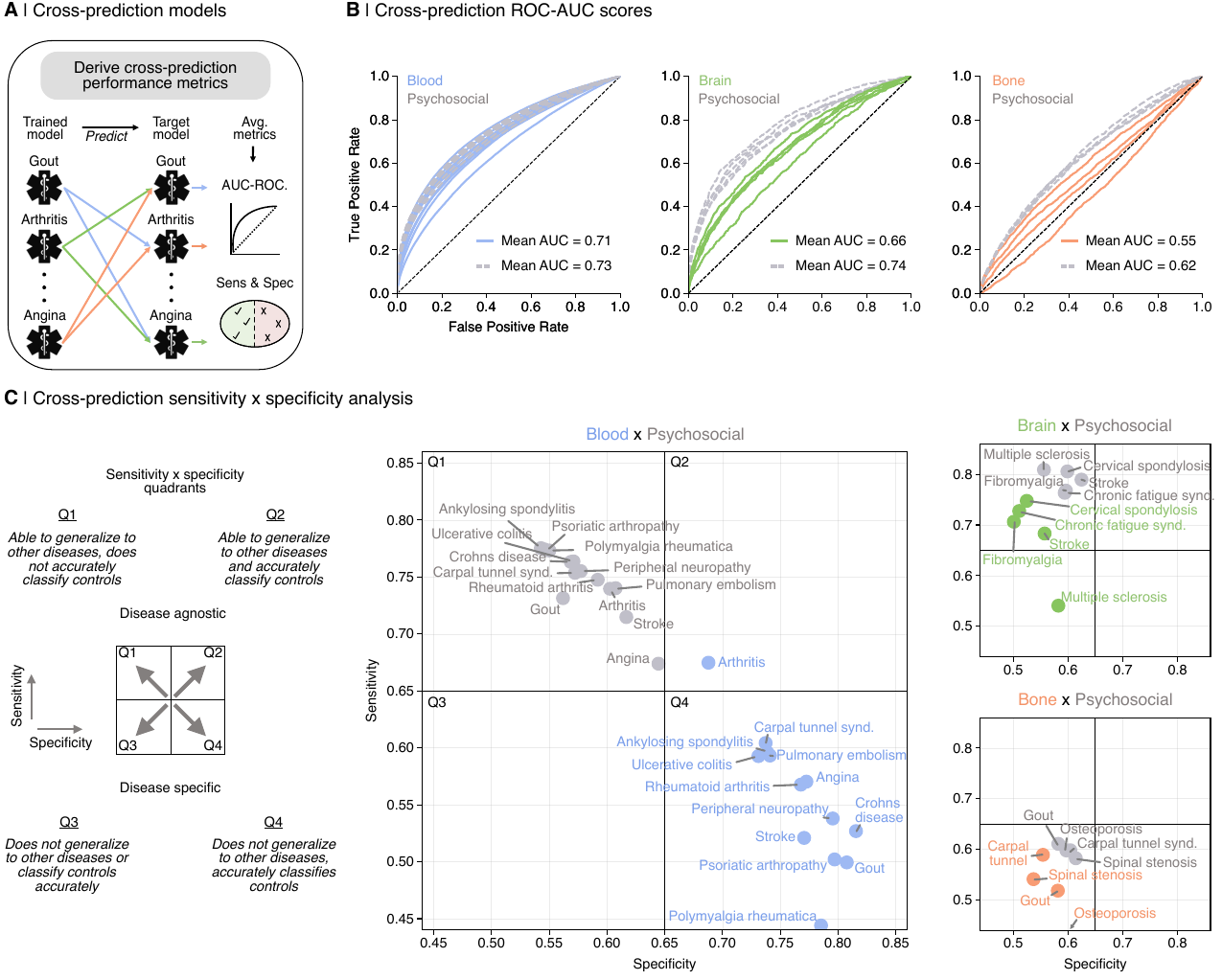

**Extended Data Figure 5: Biological and psychosocial cross-prediction models for pain diagnoses. A.** Models trained on specific diagnoses were evaluated for their ability to predict other diagnoses, with average performance metrics (ROC-AUC, sensitivity, specificity) across these diagnoses. This cross-prediction analysis was conducted for the diagnoses that were most accurately classified using blood, brain, and bone modalities alongside psychosocial models. **B.** Average cross-prediction ROC-AUC curves are displayed for both biological and psychosocial models within each biological modality. **C.** Quadrant plots show the average sensitivity and specificity of cross-prediction for each diagnosis. Points within the plots are color-coded by modality and labeled by the diagnosis on which the model was trained. Here, sensitivity measures the model's accuracy in detecting untrained diagnoses, while specificity gauges its precision in identifying diagnosis-free controls.

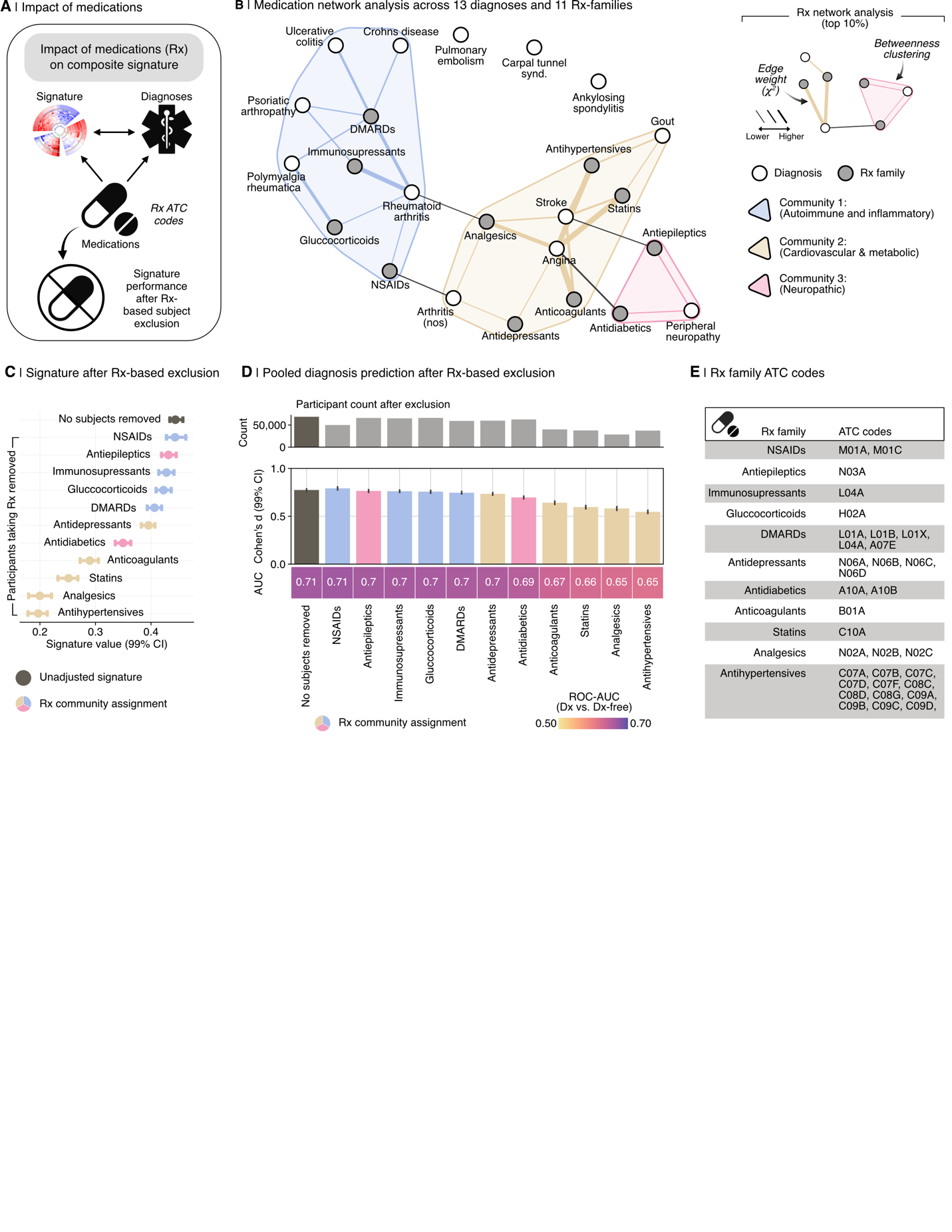

**Extended Data Figure 6: Impact of medications (Rx) on composite blood signature performance. A.** Schematic of the medication-based exclusion approach to assess medication impact on the composite signature. **B.** Network analysis using edge-betweenness clustering on chi-squared values shows medication-diagnosis communities across 13 diagnoses and 11 medication families. **C.** Adjusted composite signature values (99% CI from 1,000 bootstrap samples) for diagnosed participants, excluding those taking specific medications. **D.** The composite signature's diagnostic performance, after medication-based exclusion, in classifying 13 diagnoses is depicted using Cohen’s d and ROC-AUC, comparing diagnosed individuals to those diagnosis-free. Error bars represent the 99% confidence interval, estimated from 1,000 bootstrap samples **E.** Medication (Rx) families are organized by their corresponding Anatomical Therapeutic Chemical (ATC) classification codes and displayed in a table format.

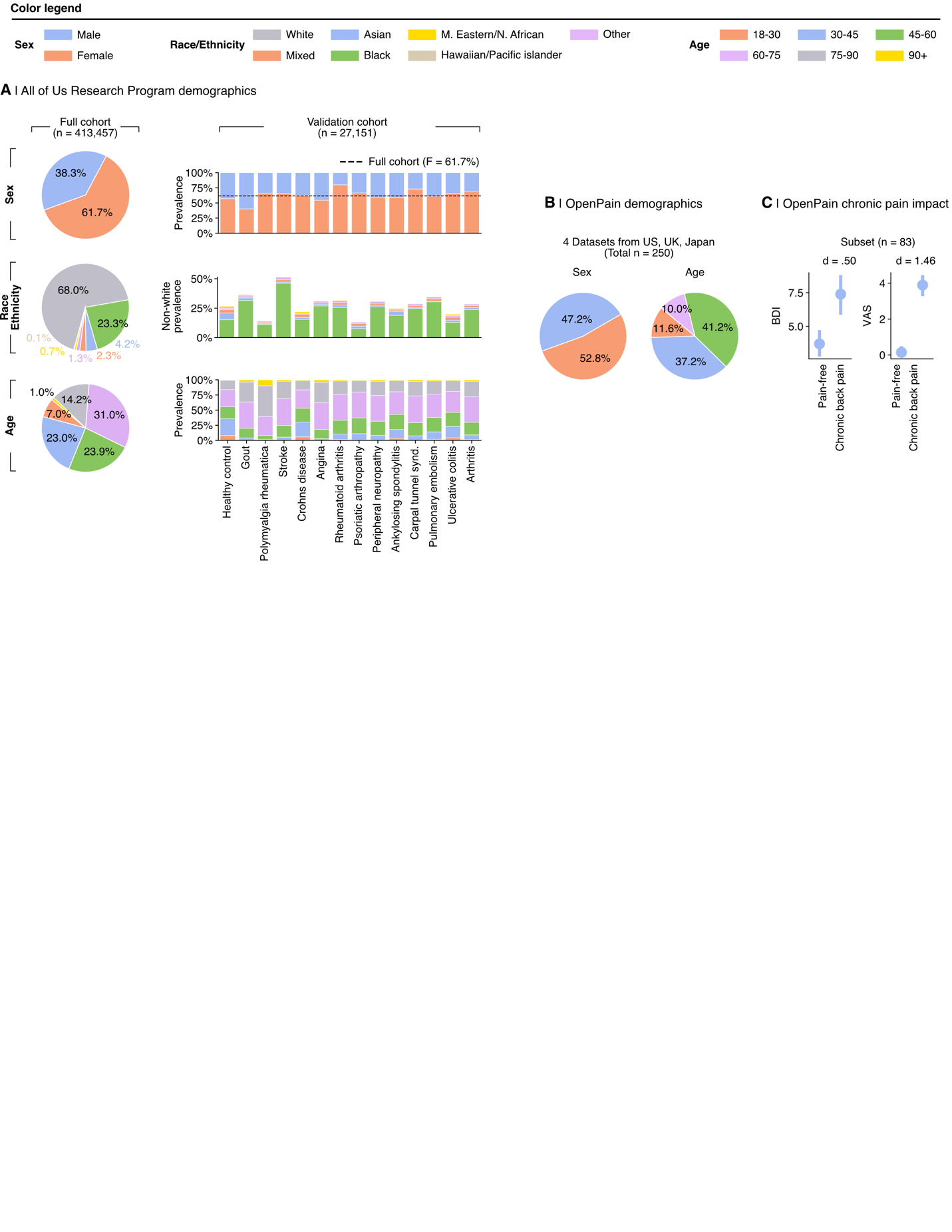

**Extended Data Figure 7: Validation datasets demographics.** Top: Color legend indicates the categories for the three demographic dimensions analyzed in each validation dataset (sex, race/ethnicity, and age). **A,** Demographics for the All of Us Research Program (AoU) are shown with pie charts for the entire cohort and stacked barplots for the subset of participants used for validation of the composite blood signature, categorized by each diagnosis and the healthy control group. Demographics are similarly depicted for the OpenPain datasets **B.** Ethnicity/race data were not available for the OpenPain datasets. **C,** Cohen’s d analysis shows the effect size of pain impact comparing patient and control groups within a subset of OpenPain, measured by the Brief Depression Inventory (BDI) and Visual Analogue Scale (VAS) for pain intensity in the last 24 hours.

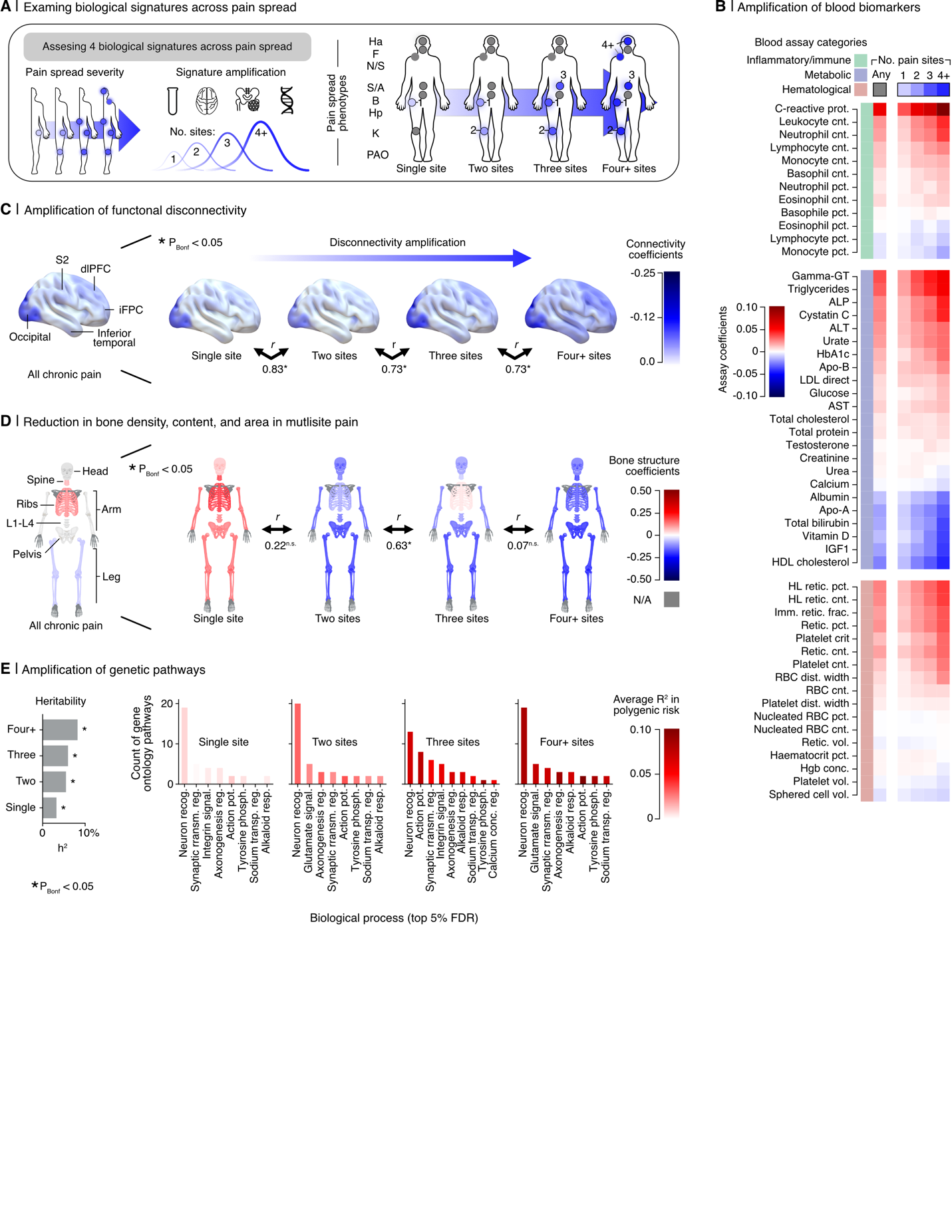

**Extended Data Figure 8: Biological pattern amplification in chronic pain spread. A.** Schematic of chronic pain spreading, quantified by the total number of chronic pain sites, alongside the investigated biological modalities: blood, brain, bone, and genetics. Biological patterns, represented by structure coefficients, associated with pain spread severity were derived from models distinguishing between levels of pain spread severity, ranging from low (one chronic pain site versus pain-free) to high (four or more chronic pain sites versus pain-free). **B.** Structure coefficients from models are shown for each level of pain spread, ordered by severity and segmented by subcategory. Additionally, coefficients for a generalized chronic pain model, encompassing any number of pain sites, are also shown. **C.** Cortical surface renderings visualize resting functional connectivity, thresholded to highlight the top 25% of structure coefficients, which represent the sum of dynamic conditional correlation across brain parcels. Arrows interlinking the cortical renderings depict the association (two-sided Pearson correlation, all *P* < 0.05 bonferonni corrected) between the complete unthresholded vectors of structure coefficients (parcel to parcel connectivity) for adjacent levels of pain spread. **D.** Skeletal body maps show bone segments colored based on structure coefficients for each assessed bone system, using DXA-derived bone density, mineral content, and area estimates. For each spreading level, averages of these three estimates for each bone system (e.g., head, spine, leg) are depicted on the maps. **E.** Heritability estimates derived from polygenic risk scores (PRS) are shown. The top 5% of FDR-corrected gene ontology pathways for each PRS are tallied and organized by biological process. The corresponding bars are shaded based on the average explained variance (R²) across pathways within each biological process.
